# Localizing at-risk and early psychosis populations along data-driven clinical, cognitive, and neuroanatomical spectra

**DOI:** 10.64898/2026.03.04.26347618

**Authors:** Matthew Danyluik, Joseph Ghanem, Saashi A. Bedford, Samantha Aversa, Alice Leclercq, Ferdousa Ibrahim, Felicia Proteau-Fortin, Joelle Eid, Matthew Morvan, Maia Turner, Sophie Piergentili, Francisco Reyes-Madrigal, Camilo de la Fuente-Sandoval, Nicholas R. Livingston, Gemma Modinos, Ridha Joober, Martin Lepage, Jai L. Shah, Yasser Iturria-Medina, M. Mallar Chakravarty

## Abstract

**Background and Hypothesis:** Psychotic disorders are thought to capture the extreme end of a psychiatric continuum, with less afflicted populations including the non-help-seeking familial high-risk state (FHR), the help-seeking clinical high-risk state (CHR), and first episode psychosis (FEP). However, we lack comprehensive analyses capturing the diversity of clinical, cognitive, functional, and neuroanatomical aberrations across all three groups, limiting our understanding of how psychosis-linked phenotypes vary in the broader scope of psychopathology.

**Study Design:** We leveraged a sample of 70 FEP, 40 CHR, 43 FHR, and 41 healthy participants recruited from the same catchment area, profiled with a cross-sectional and longitudinal multimodal battery.

**Study Results:** Using an exploratory factor analysis to treat our clinical dataset as a multidimensional spectrum, we saw that CHR expressed the most depression/anxiety symptoms, FEP endorsed the most negative/cognitive/functioning abnormalities, and FHR were comparatively less afflicted, though FEP and CHR tended to overlap along dimensions at the individual level. Regarding neuroimaging, FEP showed cortical thickness reductions resembling those seen in schizophrenia and bipolar disorder, while no consistent anatomical pattern could be identified in FHR and CHR (including in two international CHR replication cohorts). The dominant axis of brain-behaviour covariance revealed by a partial least squares correlation linked reduced cortical thickness to elevated negative and cognitive symptoms, a pattern seen most prominently in FEP. Across groups, negative and cognitive symptoms predicted lower functioning at follow-up.

**Conclusions:** CHR and FEP were defined by distinct yet overlapping clinical profiles, while neuroanatomical and joint clinical-neuroanatomical abnormalities were primarily seen in FEP.

## 1. Introduction

Psychotic disorders are thought to represent severe and chronic forms of mental health problems distributed across a much broader population^1–3^, which often evolve and intensify over time before an individual is eligible to receive a formal diagnosis and corresponding care^4–6^. In turn, clinical staging has emerged as a framework which aims to identify where an individual falls on the broader continuum of a psychiatric condition, ideally characterizing both active symptoms and the risk of clinical progression before intensive treatment is required^7–9^. In the context of psychosis, staging models typically capture familial high-risk (FHR, first-degree relatives of psychosis patients), clinical high-risk (CHR, help-seeking with subthreshold symptoms), and first episode psychosis (FEP, clinically relevant symptoms which are not yet chronic) populations, which are characterized by stepwise increases in both psychosis symptom severity and the risk of developing a chronic condition^7,10,11^.

While psychotic disorders are primarily defined by persistent psychosis symptoms, they are associated with a range of other phenomena, including comorbidities like depression and anxiety^12,13^, stable cognitive impairments^14,15^, difficulties with social and occupational functioning^16^, and widespread cortical thickness reductions^17^. Importantly, these phenotypes can also be seen to some extent in at-risk populations: FHR are characterized by subtle cognitive impairments^18,19^ and elevated risk for a range of mental health conditions^20^, while many who meet CHR criteria endorse high levels of depression and anxiety, alongside low levels of functioning^21–23^. Further, FEP is associated with cognitive impairments which can be observed upwards of decades before a first episode^24,25^ and cortical thickness reductions which can be seen as symptoms progress^26^.

However, the literature has not yet been informed by a direct, in-depth comparison of FHR, CHR, and FEP subject to clinical, cognitive, functional, and neuroanatomical profiling – perhaps because of the challenges associated with establishing infrastructure for each group in the same clinical setting. As such, we lack an understanding of which markers of psychiatric burden (cognitive impairments, schizophrenia-like cortical thickness reductions, etc.) best differentiate each group when explicitly weighed against one another. Further, we lack multimodal analyses taking advantage of the covariance between clinical and neurobiological markers across all three populations, which would provide a data-driven assessment of where each group falls along psychiatric spectra defined independently of clinical status.

Owing to the joint clinical-research infrastructure at the Douglas Mental Health University Institute in Montreal, which includes FEP and CHR services offered in the same building and a strong family engagement program, we performed a comprehensive analysis of FHR, CHR, and FEP participants profiled with a cross-sectional and longitudinal clinical, cognitive, functional, and neuroimaging battery. We leveraged a range of analytical tools to assess how each group varied along the multidimensional behavioural and neurobiological spectra present in our sample, evaluating whether at-risk and early psychosis populations occupy distinct or overlapping positions in the broader scope of psychopathology.

## 2. Methods

### 2.1. Sample

We recruited 194 participants aged 14-35 through clients and their families at the sister Prevention and Early Intervention for Psychosis (PEPP)^27^ and Clinic for the Assessment of Youth at Risk (CAYR)^28^ services, located at the Douglas Mental Health University Institute in Montreal, Canada. The sample included 70 participants diagnosed with affective or nonaffective first episode psychosis, 40 at clinical high-risk for psychosis, 43 at familial high-risk for psychosis, and 41 healthy controls. FEP and CHR participants were help-seeking and offered clinical and psychosocial treatment for up to 2 years following program entry, including individual and group cognitive behavioural therapy, social and physical activity programs, a family psychoeducation program, antidepressants, anxiolytics, and antipsychotics (the latter were available to FEP patients only). Treatment protocols for both groups are summarized in greater detail elsewhere^27,28^. Meanwhile, the FHR group included siblings of FEP patients recruited through PEPP’s family outreach program who were not seeking help for any mental health troubles (FHR participants were not necessarily siblings of the FEP participants included in this study). FEP diagnoses were determined using the Structured Clinical Interview for the Diagnostic and Statistical Manual of Mental Disorders (DSM) IV^29^, while CHR status was defined using the Structured Interview for Psychosis Risk Syndromes^30^. All assessments for the study, described below, were repeated at 6, 12, and 18 months following the baseline evaluation. Across the sample, exclusion criteria included IQ below 70, a history of brain trauma, and exclusively substance-induced psychosis. In addition to these criteria, HC were excluded if they reported a history of learning disabilities, history of any DSM diagnosis, family history of psychosis, or history of substance abuse. All participants or their guardians provided informed written consent for the use of their data for analysis. The study was approved by the Research Review Office of the Centre intégré universitaire de santé et de services sociaux du Centre-Ouest-de-l’Île-de-Montréal.

### 2.2. Clinical, cognitive, and functional assessments

FHR, CHR, and FEP participants underwent a comprehensive clinical and cognitive battery administered by trained research assistants. Evaluations included the 6-item Positive and Negative Syndrome Scale (PANSS-6)^31^, Scale for the Assessment of Positive Symptoms (SAPS)^32^, Scale for the Assessment of Negative Symptoms (SANS)^33^, Structured Interview for Psychosis Risk Syndromes (SIPS)^30^, Calgary Depression Scale for Schizophrenia (CDS)^34^, Global Assessment of Functioning (GAF)^35^, and Social and Occupational Functioning Assessment Scale (SOFAS)^36^. Cognition was assessed for all participants, including controls, using the computerized 7-domain Cogstate Research Battery (with composite scores for each domain z-scored relative to a normative population)^37,38^ and Wechsler Memory Scale-IV (WMS) (including logical memory and visual reproduction subtests, which were age-scaled)^39^. All participants also completed several symptom self-assessments, including the Beck Anxiety Inventory (BAI)^40^, 21-item Depression, Anxiety, and Stress Scale (DASS-21)^41^, Quick Inventory of Depressive Symptomatology (QIDS)^42^, Social Phobia Inventory (SPIN)^43^, Beck Cognitive Insight Scale (BCIS)^44^, and Young Mania Rating Scale (YMRS)^45^. Note that the SIPS and YMRS were only administered to the high-risk groups.

### 2.3. Statistical analysis

For each assessment, participant scores were summed across each item (or averaged for the Cogstate^37^ and subtracted from each other for the BCIS^44^) and one-way ANOVAs were used to assess group differences in summary scores after residualizing for age and sex. In cases where only two groups were assessed, a two-tailed t-test was performed instead. To correct for multiple comparisons, we applied a Holm-Bonferroni adjustment to the *p*-values from the t-tests and ANOVAs, using an alpha value of 0.05 to determine statistical significance^46^. Group differences contributing to significant differences at the omnibus level were then assessed with pairwise two-tailed t-tests, which were also corrected with the Holm-Bonferroni method. Each comparison was only performed across participants with a complete set of observations for a given scale.

### 2.4. Factor analysis

Next, we treated our clinical dataset as a multidimensional spectrum and evaluated how each group varied along its corresponding axes. We performed an exploratory factor analysis across the 153 available FHR, CHR, and FEP participants using summary scale measures from the assessments administered to all three groups (most symptoms were not evaluated in the 41 healthy controls, who were excluded from this analysis). We followed a two-stage data imputation procedure, depending on whether participants were missing either specific items or all observations for a given scale. First, missing items were replaced with the participant’s average score on the item’s scale before calculating summary scores. Then, we used the *mifa* package in R to generate 20 datasets using multiple imputation to infer missing values at the summary scale level^47^. In addition to other symptom scores, multiple imputation was informed by age, sex, and group. Imputed values were truncated at the maximum or minimum observed in the original data. Factor solutions were then derived from a Pearson’s correlation matrix pooled across the 20 imputed datasets^47^. We tested solutions with between 2 and 6 factors, applying an oblique rotation (oblimin) to each, allowing for correlations between factors and aiding interpretability^48^. We then chose the most granular solution^49^ which minimized both the number of weakly extracted factors (factors with less than 3 items with pattern loadings > 0.5) and weakly loaded items (items with pattern loadings < 0.3 on each factor)^50^.

Factor score determinacy was calculated as the multiple correlation between each factor and its optimal linear combination of observed variables^51^. Factor scores were derived using the ten Berge method, which aims for scores to correlate to the same degree as the rotated factors^51^. Note that factor scores were calculated once per imputed dataset, then averaged across datasets to give one set of scores per participant. Scores were then compared between groups using one-way ANOVAs and post-hoc Tukey’s tests to infer pairwise differences contributing to significant effects at the omnibus level. Each factor score was also used as a univariate classifier to predict group membership at the individual level, with classifier success quantified as the area under the curve (AUC) of the receiver operating characteristic curve^52^, and confidence intervals calculated using DeLong’s method^53^. We performed two sensitivity analyses: we repeated factor score group comparisons after covarying for age and sex and repeated the analysis in a subset of 104 participants who did not require multiple imputation.

Lastly, to evaluate how symptom burden evolved across follow-up visits, we generated longitudinal factor scores by applying the loadings from our chosen baseline factor solution to symptom sums from all available timepoints. 153, 105, 82, and 68 participants were available at timepoints 1, 2, 3, and 4, respectively. Missing observations were imputed across 20 multilevel multiple imputation runs using the 2l.pan approach from the *mice* package in R. Linear mixed effects models with FHR as the reference group were then used to predict factor score from group, timepoint, and their interaction, while covarying for age and sex and allowing for random intercepts by participant. Models were implemented separately within each imputed dataset and statistics were pooled via Rubin’s rules^54^. *P*-values from the interaction terms were corrected with the Holm-Bonferroni method. To confirm that symptom trajectories did not differ based on the number of available timepoints, we implemented pattern-mixture models^55^ which included an additional interaction term between time and number of visits. Finally, we assessed the impact of antipsychotic medication dosage (chlorpromazine equivalent, mg) on symptom trajectories in the FEP group. We implemented linear mixed effects models with a time-varying covariate approach^56^ to predict longitudinal factor scores from a participant’s deviation from their average medication dosage across time, covarying for average dosage, days since baseline scan, baseline age, and sex, while allowing for random intercepts by participant (66, 44, 31, and 25 FEP participants were available by timepoint for this analysis).

### 2.5. Longitudinal analysis

To evaluate which baseline symptoms best predicted future functioning, we leveraged a subset of FHR, CHR, and FEP participants with GAF assessments performed at both baseline and at least one follow-up timepoint. 105, 80, and 67 observations were available at timepoints 2, 3, and 4, respectively. For each assessment administered to all three groups, we implemented linear mixed effects models predicting future GAF from baseline summary scores, covarying for baseline GAF, age, sex, group, and timepoint while allowing for random intercepts by participant. The analysis was performed separately in each of the 20 imputed datasets used for the factor analysis above, with model results pooled across runs following Rubin’s rules^54^.

### 2.6. Neuroimaging analysis

Participants underwent a multimodal neuroimaging protocol, with exclusion criteria including claustrophobia, pregnancy, metal in any part of the body, and motor movement disorders. Here, we analyzed T1-weighted images (MPRAGE, 1 mm^3^) acquired for 190 participants with a 3T Siemens MAGNETOM Prisma Fit scanner. All images underwent manual quality control for motion artifacts following in-house guidelines^57^. Seven participants failed quality control and were excluded from all neuroimaging analyses. Five participants failed processing below, giving a total of 178 participants available for analysis.

Images were processed using FreeSurfer 7.1.0^58^ following the Enhancing Neuroimaging Genetics for Meta-Analysis (ENIGMA) FreeSurfer Protocol (https://github.com/ENIGMA-git/ENIGMA-FreeSurfer-protocol), giving cortical thickness estimates in the 68 regions of the Desikan-Killiany atlas^59^. For each group (FHR, CHR, FEP), region-wise linear models were used to calculate the effect size (Hedges’ g) of group differences in cortical thickness relative to healthy controls while covarying for age, sex, and intracranial volume. For exploratory inference, maps were thresholded at a lenient 10% false discovery rate threshold. To assess whether antipsychotics impacted our cortical thickness estimates in the FEP group, we leveraged the same time-varying covariate approach^56^ described above to predict regional cortical thickness from a participant’s deviation from their average medication dosage, covarying for average dosage, days since baseline scan, baseline age, and sex, while allowing for random intercepts by participant (60, 41, 29, and 23 FEP participants were available by timepoint for this analysis).

Using cortical thickness as an outcome measure allowed us to evaluate whether each group’s anatomical pattern resembled those seen in other common disorders. As such, we spatially correlated each unthresholded cortical thickness effect size map with corresponding maps for 8 psychiatric, developmental, and neurological conditions provided by the *ENIGMA* toolbox^60^, where each map was derived from a meta-analysis performed across upwards of thousands of images contributed by researchers worldwide to ENIGMA data sharing consortia. At the time of the analysis, case-control maps were available for 22q11.2 deletion syndrome, attention-deficit hyperactivity disorder (ADHD), autism spectrum disorder (ASD), bipolar disorder, epilepsy, major depressive disorder, obsessive-compulsive disorder (OCD), and schizophrenia. As 22q11.2 deletion syndrome is a major risk factor for psychotic disorders like schizophrenia^61^, we also included a subset of those with 22q11.2 deletion syndrome who were psychosis-positive. Since multiple maps were available for each disorder, we limited our analysis to those taken from adult samples and corrected for intracranial volume where possible (note that the latter was not the case for schizophrenia and depression). All spatial correlations were performed using the *neuromaps* toolbox^62^, where significance was assessed using permutation-based Alexander-Bloch spin tests robust to the joint spatial autocorrelation between each pair of maps^63^. A Holm-Bonferroni correction was applied to the set of *p*-values from this procedure.

### 2.7. Joint clinical-anatomical analysis

Finally, we aimed to define where each group fell on a joint clinical-anatomical continuum capturing the symptoms which most covaried with cortical thickness across the sample. We leveraged the partial least squares (PLS) correlation, a data-driven technique used to find the weighted combinations of variables from two modalities which maximally covary with each other, referred to as latent variables (LVs)^64^. Specifically, we used PLS to decompose regional cortical thickness estimates and summary scale observations into a series of clinical-anatomical LVs, with the analysis performed across 140 FHR, CHR, and FEP participants with valid neuroimaging data (i.e., the 178 participants included in the neuroimaging analysis above, excluding controls). As it was unclear how our evaluation metrics (see below) could be pooled together across multiple imputation runs, for our clinical dataset, we opted to average together the 20 imputed datasets used for our factor analysis described above. Correspondingly, as a sensitivity analysis, we re-ran PLS in a subset of 99 participants with complete neuroimaging and summary scale-level clinical data.

While PLS returns a set of LVs explaining independent covariance patterns in a sample, we opted to only interpret the first LV, as it could be directly understood as the strongest source of clinical-anatomical covariance in the sample^65^. The contribution of each brain variable to LV1 was captured with a bootstrap ratio, or the ratio between the mean and standard deviation of its weight across 1000 PLS solutions derived from bootstrap-resampled datasets^66^. The contribution of each clinical variable was represented by its loading, or the correlation between the variable and the LV across subjects, with 95% confidence intervals estimated from the same bootstrap resampling procedure^67^. LV scores, calculated after standardizing each input variable, captured the extent to which each participant expressed both the clinical and the anatomical component of the LV. Score correlations were compared between groups using Fisher’s z-test. Scores on either domain were also z-scored and summed to create composite scores measuring joint clinical-anatomical burden. Clinical, anatomical, and combined scores were compared between groups using one-way ANOVAs and post-hoc Tukey’s tests. Lastly, we also assessed whether our pattern of group differences was robust to including age and sex as covariates, as in our factor analysis.

## 3. Results

### 3.1. Sample demographics

Sample demographics are summarized in *Table 1*. CHR was significantly younger than the other groups (*Supplementary Figure 1*), while the control and FEP groups included more males than females. FEP and CHR were both help-seeking, though the time between admission for care and assessments for this study did not significantly differ between the groups and was unrelated to positive and negative symptom scores (*Supplementary Figure 2*). Most of the FEP group was treated with antipsychotic medication, while a subset (*N* = 6) of CHR participants were prescribed antipsychotics at dosages typical for anxiety or sleep problems rather than mood regulation or psychosis. FEP participants included those diagnosed with a nonaffective (schizophrenia, schizoaffective, psychosis not otherwise specified) or affective (bipolar, depressive) psychotic disorder, while most in the CHR group met the criteria for attenuated positive symptom syndrome (*Supplementary Table 1*). Only one CHR participant was known to convert to FEP at 6-, 12-, or 18-month follow-up according to SIPS criteria, though fewer than half of participants returned for all 4 timepoints (*Supplementary Table 2*). Additional sample characteristics, including self-reported medication and substance use, are shown in *Supplementary Tables 3-4*.

**Table 1:** sample demographics. The first 4 columns show group-level summary statistics, while the final column shows the results of an uncorrected one-way ANOVA or chi-squared test assessing group differences in the statistic indicated. Note that 6 CHR participants reported taking antipsychotics, which were not prescribed at dosages intended to manage psychosis symptoms.

|  | HC (N = 41) | FHR (N = 43) | CHR (N = 40) | FEP (N = 70) | Statistic |
| --- | --- | --- | --- | --- | --- |
| **Age** (mean, std) | 24.22 (4.38) | 25.47 (5.69) | 20.75 (4.96) | 25.10 (4.59) | $F(3,190) = 8.19$,<br>$p < 0.001$ |
| **Male sex** (%) | 68.3 | 46.5 | 47.5 | 65.7 | $\chi^2(3,194) = 7.64$,<br>$p = 0.054$ |
| **White race** (%) | 63.4 | 48.8 | 64.1 | 48.6 | $\chi^2(3,193) = 4.31$,<br>$p = 0.230$ |
| **Years of education**<br>(mean, std) | 13.51 (1.59) | 13.35 (2.76) | 10.94 (2.32) | 12.34 (2.37) | $F(3,189) = 10.37$,<br>$p < 0.001$ |
| **Chlorpromazine equivalent, mg**<br>(mean, std) |  |  | 61.72 (25.01) | 334.62 (228.03) |  |
| **Days since program entry**<br>(mean, std) | | | 69.15 (56.93) | 60.89 (32.24) | $t(90) = 0.87$,<br>$p = 0.388$ |
| **Neuroimaging quality control pass rate** (%) | 100 | 97.6 | 92.3 | 95.7 | $\chi^2(3,190) = 3.58$,<br>$p = 0.310$ |

### 3.2. Clinical, cognitive, and functional assessments

First, we evaluated group differences in summary scores from our battery of 15 clinical, cognitive, and functional assessments, with results shown in *Table 2*. After correcting for age and sex, CHR and FEP expressed positive and negative psychosis symptoms to the same degree, alongside equally low levels of global and social/occupational functioning. Meanwhile, cognitive performance was significantly lower in FEP relative to FHR, CHR, and HC. CHR tended to express more depression and anxiety symptoms than the other groups, but these comparisons did not survive a multiple comparisons correction. FHR did not significantly differ from controls on any scale where the latter was assessed. However, uncorrected *p*-values comparing group means on each item revealed trends towards lower cognition and greater levels of depression/anxiety in FHR relative to controls on some domains (for visualizations of item-level scores across all groups, see *Supplementary Figures 3-6*).

**Table 2:** clinical, cognitive, and functional evaluations performed across the sample. The first 4 columns show the mean and standard deviation of summary scores for the assessment and group indicated, while the last column shows the results of an omnibus one-way ANOVA or two-tailed t-test comparing summary scores between groups after residualizing scores for age and sex. Bolded comparisons were significant after Holm-Bonferroni correction. Group comparisons deemed significant after multiple comparisons correction are indicated below each omnibus test result. Note that comparisons were only performed across participants with complete data for a given assessment.

|  | HC (N = 41) | FHR (N = 43) | CHR (N = 40) | FEP (N = 70) | Statistic |
| --- | --- | --- | --- | --- | --- |
| **PANSS-6**<br>Positive and Negative<br>Syndrome Scale - 6 items<br>(Positive) | | 3.60 (1.27) | 7.18 (2.98) | 6.47 (3.46) | $F(2,137) = 16.31$,<br>$p < 0.001$<br>FEP, CHR > FHR |
| **PANSS-6**<br>Positive and Negative<br>Syndrome Scale - 6 items<br>(Negative) | | 3.55 (1.24) | 6.51 (3.51) | 7.36 (3.77) | $F(2,137) = 16.15$,<br>$p < 0.001$<br>FEP, CHR > FHR |
| **SANS**<br>Scale for the Assessment<br>of Negative Symptoms | | 1.29 (2.05) | 6.54 (3.82) | 7.60 (4.74) | $F(2,145) = 30.98$,<br>$p < 0.001$<br>FEP, CHR > FHR |
| **SAPS**<br>Scale for the Assessment<br>of Positive Symptoms | | 0.88 (1.50) | 4.77 (3.09) | 4.62 (4.14) | $F(2,147) = 16.98$,<br>$p < 0.001$<br>FEP, CHR > FHR |
| **SIPS**<br>Structured Interview for<br>Psychosis Risk Syndromes | | 7.00 (7.74) | 35.53 (12.50) | | $t(78) = 9.15$,<br>$p < 0.001$ |
| **YMRS**<br>Young Mania Rating Scale | | 1.02 (2.10) | 4.35 (3.73) | | $t(78) = 3.19$,<br>$p = 0.002$ |
| **CDS**<br>Calgary Depression Scale<br>for Schizophrenia | | 1.14 (3.06) | 5.90 (4.54) | 4.15 (4.00) | $F(2,146) = 13.43$,<br>$p < 0.001$<br>FEP, CHR > FHR |
| **QIDS**<br>Quick Inventory of<br>Depressive<br>Symptomatology | 3.41 (2.48) | 6.58 (5.08) | 14.74 (6.13) | 10.30 (4.39) | $F(3,118) = 23.03$,<br>$p < 0.001$<br>FEP, CHR ><br>FHR, HC |
| **DASS-21**<br>Depression, Anxiety, and<br>Stress Scale - 21 items | 16.00 (13.59) | 24.94 (22.25) | 67.31 (30.00) | 43.04 (27.08) | $F(3,140) = 20.75$,<br>$p < 0.001$<br>FEP, CHR ><br>FHR, HC |
| **BAI**<br>Beck Anxiety Inventory | 5.74 (5.16) | 11.00 (10.85) | 23.93 (15.43) | 13.96 (9.83) | $F(3,138) = 10.86$,<br>$p < 0.001$<br>FEP, CHR ><br>FHR, HC |
| **SPIN**<br>Social Phobia Inventory | 11.04 (2.48) | 12.53 (8.15) | 27.21 (14.00) | 20.96 (15.25) | $F(3,139) = 9.61$,<br>$p < 0.001$<br>FEP, CHR ><br>FHR, HC |
| **Cogstate**<br>Computerized Research<br>Battery | -0.12 (0.51) | -0.33 (0.68) | -0.23 (0.43) | -0.74 (0.86) | $F(3,186) = 8.54$,<br>$p < 0.001$<br>FEP <<br>HC, FHR, CHR |
| **WMS**<br>Wechsler Memory Scale-IV | 41.02 (7.75) | 37.52 (9.98) | 39.61 (7.36) | 34.06 (9.11) | $F(3,185) = 6.19$,<br>**$p < 0.001$**<br>FEP <<br>HC, FHR, CHR |
| **BCIS**<br>Beck Cognitive Insight<br>Scale | 2.81 (4.61) | 4.12 (4.46) | 9.07 (5.82) | 6.58 (5.15) | $F(3,160) = 9.23$,<br>**$p < 0.001$**<br>FEP, CHR ><br>FHR, HC |
| **GAF**<br>Global Assessment of<br>Functioning | | 82.69 (11.70) | 54.08 (8.85) | 53.94 (17.09) | $F(2,147) = 54.55$,<br>**$p < 0.001$**<br>FEP, CHR < FHR |
| **SOFAS**<br>Social and Occupational<br>Functioning Assessment<br>Scale | | 83.10 (10.84) | 58.69 (10.55) | 58.22 (15.58) | $F(2,143) = 45.97$,<br>**$p < 0.001$**<br>FEP, CHR < FHR |

### 3.3. Factor analysis

Next, we aimed to derive the latent dimensions of joint clinical, cognitive, and functional variability which defined our sample, then quantify where FHR, CHR, and FEP fell along each data-driven axis. We performed an exploratory factor analysis across our psychosis spectrum groups (*N* = 153) using the battery of summary scores described above, without encoding any explicit information on group membership. We chose a factor solution with three correlated symptom profiles (*Supplementary Figure 7*) capturing, respectively, depression/anxiety; greater negative symptoms and lower cognition/functioning; and greater positive symptoms and lower functioning (*Figure 1A*, 62.8% variance explained). When comparing factor scores capturing the extent to which participants expressed each symptom profile, we observed significant omnibus effects for all three dimensions (depression-anxiety: *F*(2,150) = 24.90, *pHolm* < 0.001; negative-cognitive-functioning: *F*(2,150) = 32.90, *pHolm* < 0.001; positive-functioning: *F*(2,150) = 34.51, *pHolm* < 0.001). Post-hoc comparisons revealed stepwise increases from FHR to CHR to FEP along the negative-cognitive-functioning dimension, while CHR and FEP loaded equivalently onto the positive-functioning axis relative to FHR, and CHR expressed significantly more depression-anxiety symptoms than the other two groups (*Figure 1B*). Linear mixed effects models predicting longitudinal factor scores revealed a significant group-by-time interaction, where positive-functioning symptoms declined more quickly in FEP relative to FHR (*β* = −0.22, *pHolm* = 0.015) (*Supplementary Figure 8*). We also observed a trend towards a steeper decline in depression-anxiety symptoms in CHR relative to FHR (*β* = −0.21, *pHolm* = 0.075). Changes in medication dosage did not significantly predict changes in factor scores over time (*Supplementary Figure 9*).

**Figure 1:**
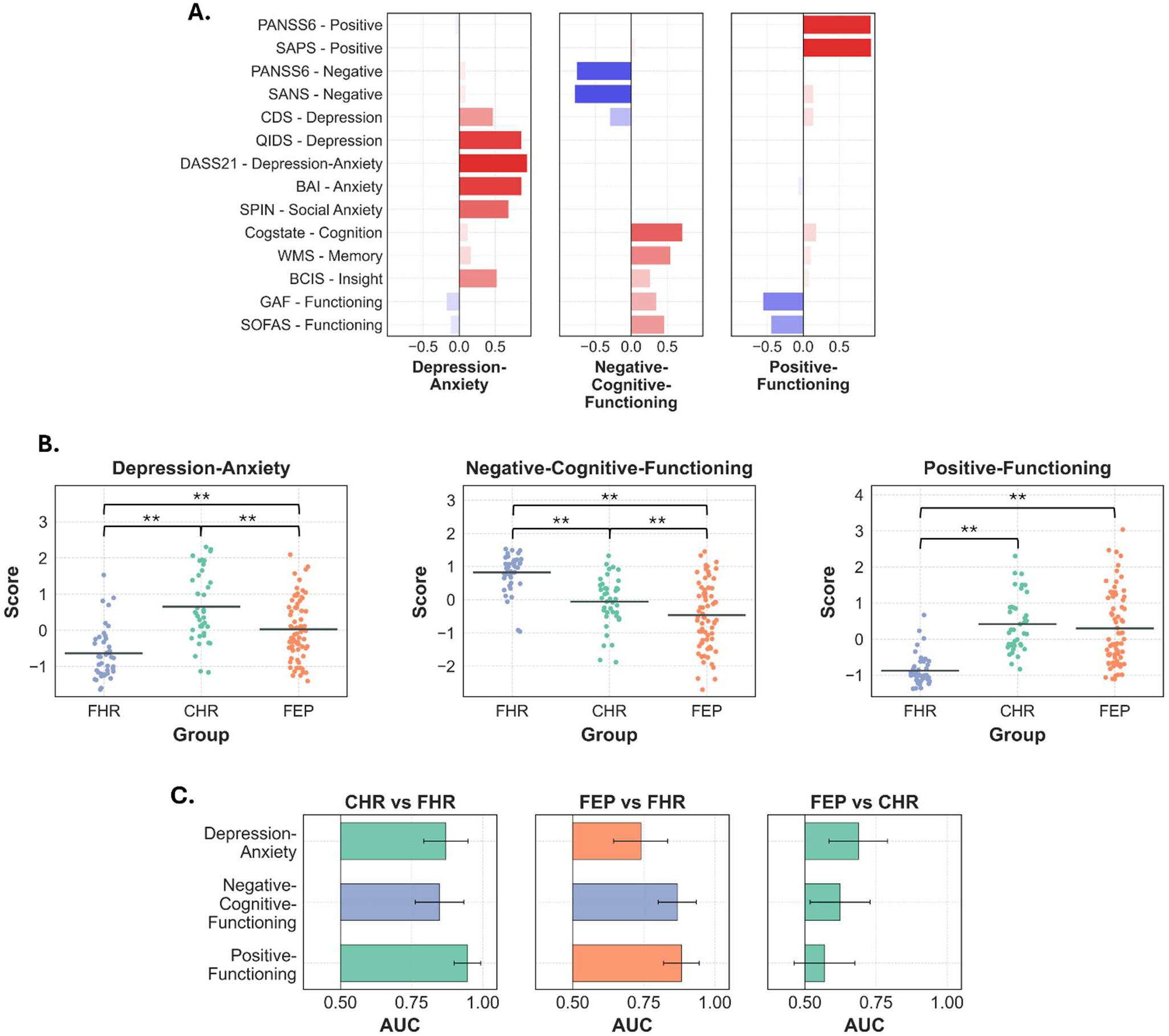
**(A)** Pattern loadings for the chosen three factor solution. **(B)** Comparing factor scores between groups, where a more positive score indicates greater expression of the symptom profile captured by the factor. CHR expressed the most depression-anxiety symptoms, while FEP showed the most negative-cognitive-functioning aberrations. FEP and CHR expressed the positive-functioning symptom profile to the same degree. \*\**p_Tukey_ < 0.05*. **(C)** AUC values quantifying the overlap between groups on each factor. CHR and FEP could only be modestly distinguished from each other relative to comparisons involving FHR. Bar colour indicates the group with the more positive mean factor score. **Alt Text:** The symptoms which are seen together across at-risk and early psychosis groups, alongside how much each individual expresses each symptom profile.

Despite the presence of significant group differences in mean factor scores, area under the curve (AUC) values quantifying the extent to which factor scores could predict group membership at the individual level indicated only modest separability of CHR and FEP along the three dimensions (0.69, 0.62, and 0.57 respectively, where 0.5 indicates complete overlap and 1.0 indicates perfect separability), while FHR was more distinguishable from the other groups (0.87, 0.85, and 0.95 relative to CHR; 0.74, 0.87, 0.88 relative to FEP) (*Figure 1C*). Note that factor scores were highly determinable and reproduced the correlation structure of the factor solution, and that variable communalities were generally high as well (*Supplementary Figure 10*). Group contrasts were largely consistent after residualizing factor scores for age and sex (*Supplementary Figure 11*) and in a subset of participants (*N* = 104) who were not missing any summary scale observations (*Supplementary Figure 12*). See *Supplementary Tables 5-6* for a summary of data requiring imputation.

### 3.4. Longitudinal analysis

Leveraging the longitudinal component of our study, we assessed which baseline symptoms best predicted future functioning, analyzing a subset of 105 FHR, CHR, and FEP participants who completed the GAF at baseline and at least one follow-up timepoint. Linear mixed effects models demonstrated that greater baseline cognition (Cogstate: *β* = 0.24, *pHolm* = 0.001) and lower baseline negative symptoms (SANS: *β* = −0.30, *pHolm* < 0.001; PANSS-6 negative: *β* = −0.27, *pHolm* = 0.001) significantly predicted greater follow-up functioning (*Supplementary Figure 13*).

### 3.5. Neuroimaging analysis

Next, we explored the neuroanatomical profiles which defined each group. Unthresholded effect size maps quantifying group differences in cortical thickness relative to healthy controls revealed a broad pattern of cortical thickness reductions in FEP which was not evident in the two high-risk groups (*Figure 2*), despite the fact that CHR and FEP were both clinically impaired relative to FHR (*Figure 1*). Three regions in the FEP map (left pars orbitalis, rostral middle frontal, and supramarginal cortex) differed from controls at an exploratory 10% false discovery rate threshold (*Supplementary Figure 14*). Group contrasts were reproducible when FHR or CHR served as the reference population (*Supplementary Figure 15*). Leveraging the longitudinal component of our study, we saw no evidence that deviations in antipsychotic dosage over time predicted corresponding deviations in cortical thickness in FEP (*Supplementary Figure 16*).

**Figure 2:**
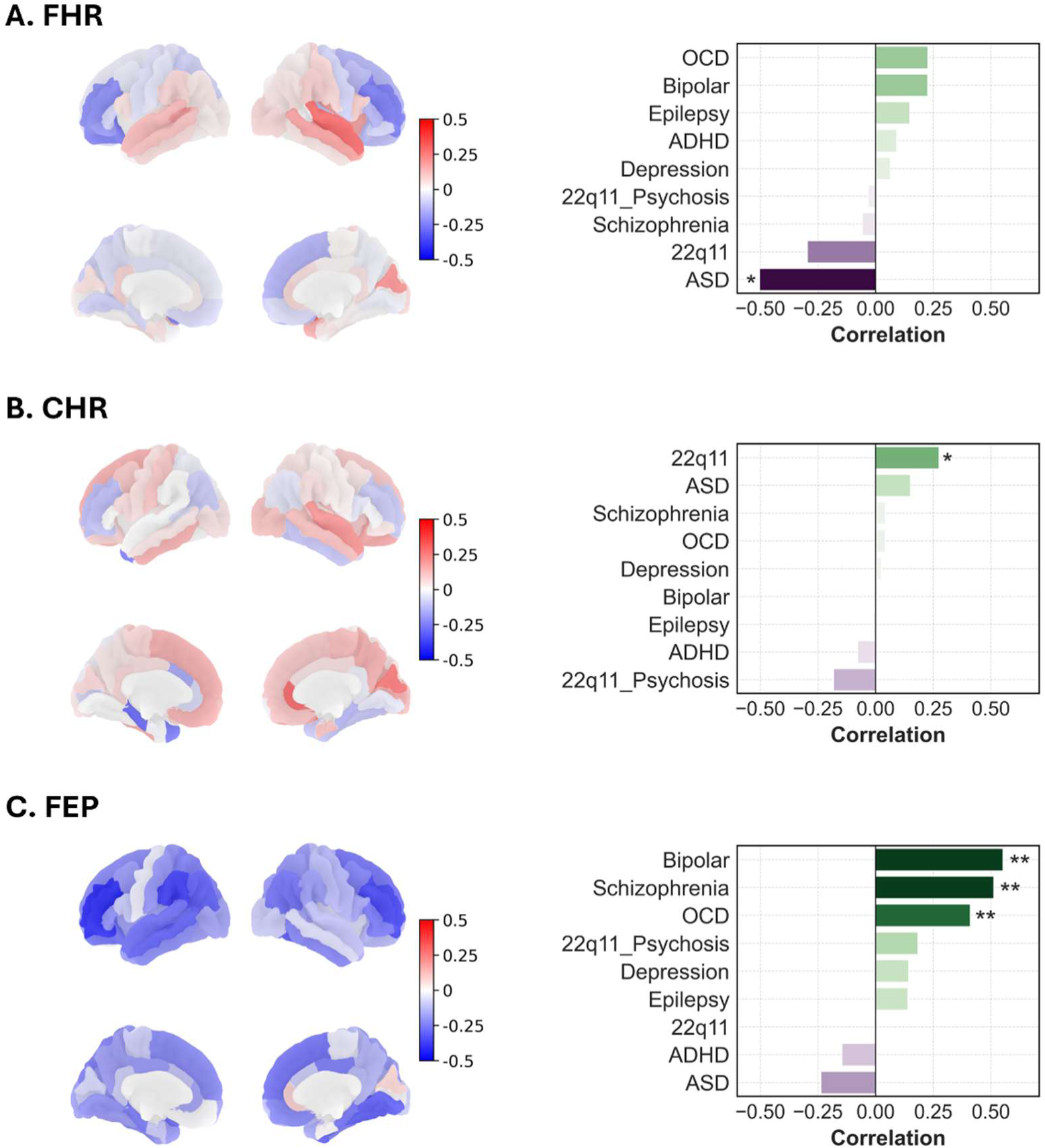
(*Left*) unthresholded regional effect of group (Hedges’ g) on cortical thickness relative to healthy controls while covarying for age, sex, and intracranial volume for FHR **(A)**, CHR **(B)**, and FEP **(C)**. FEP was characterized by broad reductions in cortical thickness relative to controls, while the pattern was much less consistent in the high-risk groups. (*Right*) Spatial correlations between each effect size map and maps for 8 psychiatric, neurological, and developmental conditions provided by the ENIGMA toolbox. FEP was the only group with a schizophrenia-like cortical thickness pattern. *\*p < 0.05, **p_Holm_ < 0.05*. **Alt Text:** The cortical thickness profiles of each at-risk and early psychosis group, alongside which disorders these maps resemble.

To qualify each group’s cortical thickness map, and determine whether each pattern of group differences captured a similar phenotype to what is typically seen in schizophrenia, we spatially correlated each cortical thickness effect size map with those derived from meta-analyses of schizophrenia and 7 other psychiatric, developmental, and neurological conditions provided by the *ENIGMA* toolbox (*Figure 2*). As expected, the FEP cortical thickness pattern strongly and significantly resembled that seen in schizophrenia (*r* = 0.51, *p* < 0.001), alongside bipolar disorder (*r* = 0.55, *p* < 0.001) and OCD (*r* = 0.41, *p* < 0.001). Conversely, neither high-risk group showed a schizophrenia-like cortical thickness pattern: the CHR map best resembled that of 22q11.2 deletion syndrome (*r* = 0.28, *p* = 0.045), a major genetic risk factor for schizophrenia and several developmental disabilities^60^, while the FHR profile was inversely related to that of autism (*r* = −0.50, *p* = 0.027), though neither comparison survived a multiple comparisons correction. Note as well that the CHR phenotype could not be replicated, even at the trend level, in two held-out international cohorts from King’s College London^68^ and the Instituto Nacional de Neurología y Neurocirugía in Mexico City^69^ (*Supplementary Figure 17,* see *Supplementary Tables 7-8* for sample characteristics), suggesting that CHR could not be described by a distinct group-level neuroanatomical profile as we observed in FEP.

### 3.6. Joint clinical-anatomical analysis

Lastly, we aimed to define where each group fell on a data-driven continuum capturing joint variability in symptoms and neuroanatomy. We performed a partial least squares (PLS) correlation across each participant’s behavioural assessments and cortical thickness measures, which returned the sets of clinical and anatomical variables which covaried regardless of group status. The strongest clinical-anatomical axis returned by PLS (49.3% covariance explained) revealed that lower cortical thickness across parietal, temporal, and cingulate regions was related to lower cognitive performance and more negative symptoms (*Figure 3A-B*). Clinical and anatomical scores capturing the extent to which each participant expressed either part of the latent component were significantly correlated across the sample (*r* = 0.271, *p* = 0.001) (*Figure 3C*). The relationship was the strongest and most significant within FEP (FEP: *r* = 0.273, *p* = 0.029; CHR: *r* = 0.173, *p* = 0.312; FHR: *r* = 0.128, *p* = 0.432), though correlations did not significantly differ from each other (*z* = 0.75, *p* = 0.449) (*Supplementary Figure 18*). Scores significantly differed between groups (anatomical: *F*(2,137) = 8.01, *p* = 0.001; clinical: *F*(2,137) = 27.50, *p* < 0.001), where FEP expressed only the anatomical, and not the clinical, pattern more than the high-risk cohorts (*Supplementary Figure 18*). Combined scores measuring joint burden on both the clinical and anatomical domains also significantly differed between groups (*F*(2,137) = 20.56, *p* < 0.001), where FEP endorsed the combined pattern to a greater degree than both CHR and FHR (*Figure 3D*). In sum, the dominant clinical-anatomical pattern in our sample appeared more prominent in FEP than in the high-risk groups. Group comparisons were consistent after residualizing scores for age and sex (*Supplementary Figure 19*), while the dominant covariance pattern was reproducible in a subset of participants who did not require data imputation (*Supplementary Figure 20*).

**Figure 3:**
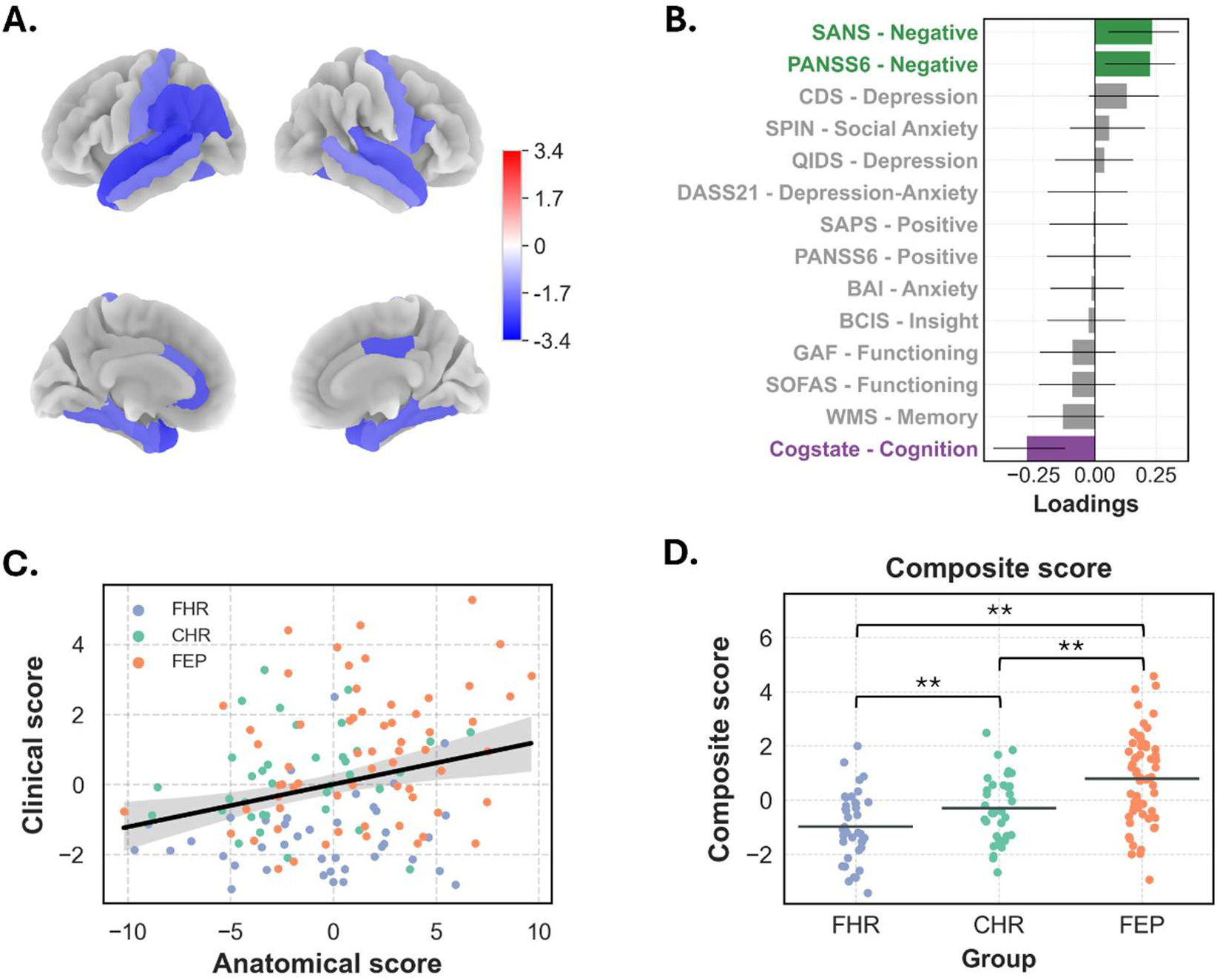
The first latent variable from the partial least squares analysis, showing the set of clinical and anatomical variables which most covaried across our sample. The latent variable captured reduced cortical thickness in temporal/parietal regions **(A)** which related to more negative symptoms and depression, as well as lower levels of cognition **(B)**. The strength of each brain variable is represented by a bootstrap ratio (interpretable as a z-score and thresholded at 1.96, the critical z-value for *p = 0.05*, accordingly), while the strength of each clinical variable is represented by a loading with bootstrapped 95% confidence intervals (see *Methods*). Scores are plotted in **(C)**, group comparisons in composite scores **(D)**, where FEP expressed the combined clinical-anatomical pattern the most. **Alt Text:** The clinical and neuroanatomical profiles which are seen together across at-risk and early psychosis groups, alongside how much each group expresses each of these patterns.

## 4. Discussion

Owing to a clinical infrastructure which allowed us to recruit and profile FHR, CHR, and FEP participants from the same catchment area, we performed a comprehensive assessment of the clinical, cognitive, functional, and neuroanatomical markers which define high-risk and first episode populations. While we observed significant mean differences between groups, we also saw overlap between FEP and CHR when analyzing where individuals fell on data-driven axes of variability. For one, at the group level, our exploratory factor analysis revealed stepwise increases in negative/cognitive/functioning abnormalities from FHR to CHR to FEP, alongside higher levels of depression/anxiety in CHR relative to FHR/FEP. Positive/functioning symptoms tended to improve in FEP over time as well. However, AUC values demonstrated that CHR and FEP overlapped along these dimensions considerably more than either group overlapped with FHR, suggesting that help-seeking individuals can express a range of symptom profiles regardless of their clinical status. Similarly, our PLS analysis demonstrated that cognitive and negative symptoms most covaried with cortical thickness across the sample, with the clinical pattern equally prominent in both CHR and FEP. In turn, our data-driven approach suggests that the clinical boundary between CHR and FEP, defined largely by positive symptom severity^7^, does not necessarily correspond to distinct breakpoints in other underlying behavioural spectra. Accordingly, our results may align with efforts to incorporate early psychosis services into more general transdiagnostic mental health infrastructure, which recognizes the phenotypic overlap seen across diagnostic categories and tailors care to each individual’s clinical, cognitive, and functional profile^8,9,11^.

Across our analyses, joint negative and cognitive symptoms related to various outcomes of interest. For one, lower cognitive performance (and negative-cognitive-functioning covariability) defined FEP at the group level, aligning with previous work suggesting that cognitive impairments are a stable, defining feature of psychosis, perhaps conferring risk well before frank symptoms emerge^24,70,71^. Greater negative symptoms and lower cognitive performance also predicted lower functioning at follow-up, generalizing previous findings in CHR^72,73^ and in FEP^74,75^ to the broader sample studied here. Lastly, as discussed, our PLS analysis showed that cognition and negative symptoms covaried with cortical thickness across our populations, extending previous “doubly multivariate” work which linked brain volume with negative symptoms/cognition^76^ and cortical thickness with cognition^77–79^ in schizophrenia alone. Given that negative and cognitive symptoms are often undertreated in clinical settings^80^, our results support ongoing efforts to better address these symptoms with novel interventions such as cognitive remediation therapy^81,82^.

Beyond capturing relative clinical overlap between FEP and CHR, our analysis also demonstrated neuroanatomical divergence in FEP relative to the high-risk populations. For one, FEP was the only group to show a pattern of cortical thickness reductions characteristic of those seen in schizophrenia, suggesting that a psychosis-like anatomical profile may only be fully observable if frank psychotic symptoms emerge^26,83^. The FEP anatomical pattern resembled that of bipolar disorder as well, perhaps reflecting the prevalence of affective psychosis in our sample and indicating that our FEP group captured a broader psychosis phenotype. Critically, FEP also expressed the joint clinical-anatomical pattern from our PLS analysis more than the other groups, a tendency which was not evident when considering the clinical component alone. As such, symptom burden and neuroanatomy may be most strongly coupled in more severely afflicted clinical populations, as suggested elsewhere in the neuroimaging literature^84,85^.

Meanwhile, despite being characterized by a broad symptom profile, CHR did not endorse any consistent pattern of cortical thickness reductions across our sample or two international replication cohorts. While a large-scale meta-analysis of the high-risk state identified small yet significant reductions in cortical thickness relative to controls^86^, it is thought that most anatomical variability in CHR is nested within normative ranges^87,88^, and that differences from controls are driven by a subset of individuals who are more likely to convert to psychosis^86,88^ or to express schizophrenia-like cognitive impairments^89^. Given that our CHR population included only one known converter and did not differ from controls regarding cognitive performance, our results suggest that it may only be feasible to identify a consistent cortical profile in high-risk populations who demonstrate more psychosis-like phenotypes. Instead, non-structural neural markers such as cerebral blood flow^90^, or measures taken from subcortical regions like the hippocampus and striatum, may be more sensitive to psychosis-like abnormalities in subthreshold states – a topic which we aim to address in future work, leveraging the multimodal neuroimaging component of our dataset.

We note several limitations of our work. First, our sample size was modest relative to large, multi-centre early psychosis cohorts, including NAPLS^91^ and PSYSCAN^92^. This point is especially relevant given the scope of our analysis, and our use of multivariate techniques in a smaller subsample which excluded controls. Many effects did not survive a multiple comparisons correction, and as such, any null results we reported cannot necessarily be interpreted as an absence of an effect. Next, we saw no differences in positive symptom scores between CHR and FEP, which may have been a product of antipsychotic treatment in the FEP group. However, it is also possible that our cross-sample assessments, which were designed to measure psychosis symptoms in clinical populations, were not as sensitive to psychopathology in the subthreshold CHR state^93^. Further, while we attempted to assess antipsychotic effects in FEP for our neuroimaging analysis, it is difficult to completely deconfound medication and psychosis severity. Reassuringly, recent work has shown that longitudinal cortical thinning is preferentially seen in FEP patients treated with placebo rather than antipsychotics^94^, and that reduced cortical thickness can be observed in never-medicated psychosis patients^95^, suggesting that neuroanatomical abnormalities are primarily symptom-related in psychosis populations.

We also acknowledge that our high-risk groups did not necessarily behave like others reported in the literature. We did not identify any significant differences in psychopathology^20,96^ or cognition^18,19,97,98^ relative to controls in our FHR group, though we may have been underpowered to do so, given that uncorrected *p*-values revealed trends towards elevated symptom burden in the sibling cohort on several cognitive and depression/anxiety domains. However, our FHR group was limited to siblings of FEP patients, while other FHR groups consist of children of those afflicted with a severe mental illness – a population where genetic factors and environmental stressors introduced while being raised by an affected parent^99,100^ may interact to increase liability for developing psychiatric symptoms. Our FHR group was also non-help seeking, and by extension perhaps captured the most resilient subset of the broader FHR construct. Meanwhile, our CHR group consisted of a small proportion of known converters relative to the expected rate of 20% after 18 months^101^, the latest follow-up observation in our study, indicating that our CHR group was not as severely afflicted as others in the literature (though less than half of CHR participants returned for all follow-up visits). Lastly, we note that since CHR was younger than the other groups in our study, any psychopathology we observed in CHR may have simply been underdeveloped relative to the other cohorts.

Together, we profiled a densely phenotyped cohort spanning three at-risk and early psychosis populations. At the group level, CHR was defined by a broad clinical profile including attenuated psychosis symptoms, depression/anxiety, and difficulties functioning, while cognitive and neuroanatomical abnormalities were more prevalent in FEP. After defining data-driven axes of clinical variability, we saw that the CHR-FEP boundary was only modestly separable relative to the boundary between either group and FHR. Meanwhile, negative and cognitive symptoms tracked lower cortical thickness, a brain-behaviour relationship which was more established in FEP than the high-risk cohorts. Our results indicate that clinical, cognitive, and functional phenotypes can discriminate between high-risk and early psychosis populations, with meaningful overlap at the individual level, while neuroanatomical burden and its relationships with behaviour can be seen more selectively in those who experienced a first episode.

## Data Availability

All data produced in the present study are available upon reasonable request to the authors.

## Acknowledgements

This work was supported by the Canadian Institutes of Health Research (CIHR); the Natural Sciences and Engineering Research Council of Canada (NSERC); le Fonds de recherche du Québec – Santé (FRQS); and the Canada First Research Excellence Fund (CFREF), awarded through the Healthy Brains, Healthy Lives (HBHL) initiative at McGill University.

