## Supplementary Materials for "Localizing at-risk and early psychosis populations along data-driven clinical, cognitive, and neuroanatomical spectra"

### 1. Demographics

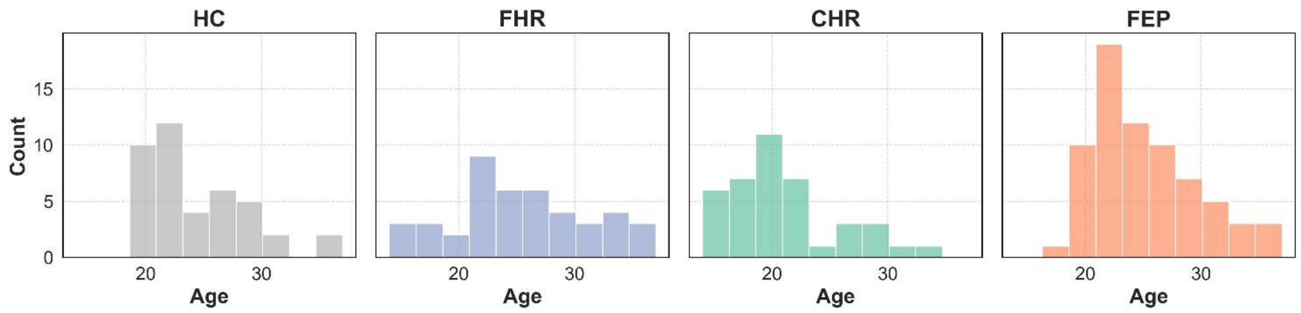


***Supplementary Figure 1****:* age distribution across the sample. Age significantly differed between groups ($F(3,190) = 8.19$, $p < 0.001$). Post-hoc Tukey's tests suggested that CHR were significantly younger than the other three groups.

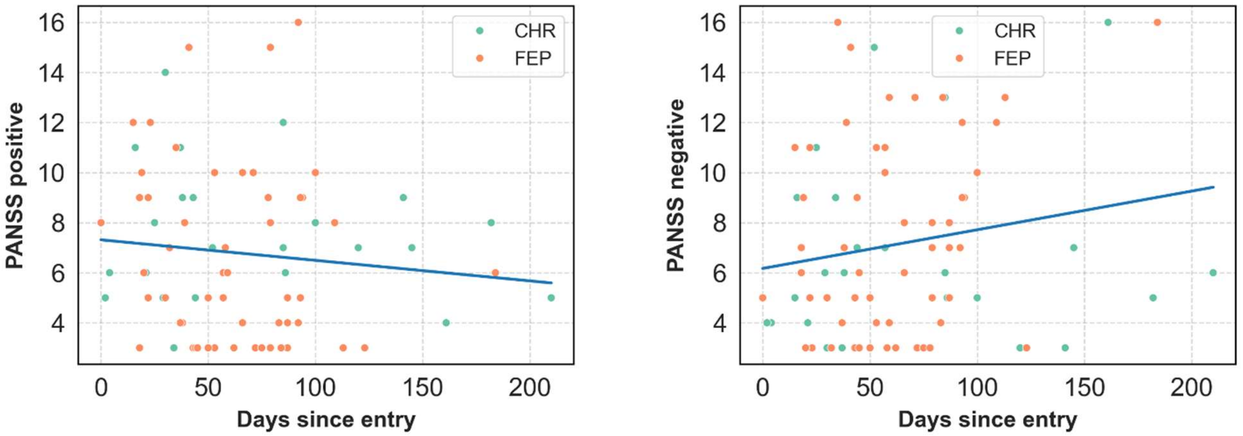


***Supplementary Figure 2****:* relationship between days since program entry and PANSS-6 positive (*Left*) and negative (*Right*) summary scores. Positive symptoms tended to decrease, $t(79) = -0.93$, $p = 0.353$, and negative symptoms tended to increase, $t(79) = 1.57$, $p = 0.121$, with time since entry, though neither comparison was significant.

| | CHR (*N* = 40) |
|---|---|
| **Attenuated Positive Symptom Syndrome (APSS)** | 28 (70%) |
| **Brief Intermittent Psychotic Syndrome (BIPS)** | 1 (2.5%) |
| **Genetic Risk and Deterioration Syndrome (GRD)** | 2 (5%) |
| **APSS + BIPS** | 4 (10%) |
| **Unknown** | 5 (12.5%) |

| | FEP (*N* = 70) |
|---|---|
| **Schizophrenia and Other Psychotic Disorders** | 33 (47.1%) |
| **Bipolar Disorders** | 12 (17.1%) |
| **Depressive Disorders** | 6 (8.6%) |
| **Personality Disorders** | 1 (1.4%) |
| **Unknown** | 18 (25.7%) |

***Supplementary Table 1:*** (*Left*) prevalence of SIPS syndromes in the CHR group. (*Right*) prevalence of DSM-IV diagnoses in the FEP group.

| | HC | FHR | CHR | FEP |
|---|---|---|---|---|
| **Baseline** | 41 (100%) | 43 (100%) | 40 (100%) | 70 (100%) |
| **6-month** | 31 (75.6%) | 26 (60.5%) | 26 (65%) | 53 (75.7%) |
| **12-month** | 27 (65.9%) | 21 (48.8%) | 21 (52.5%) | 40 (57.1%) |
| **18-month** | 23 (56.1%) | 16 (37.2%) | 15 (37.5%) | 37 (52.9%) |

***Supplementary Table 2:*** participants by group by timepoint. Fewer than half of FHR and CHR participants returned for all 4 timepoints.

| | HC (*N* = 41) | FHR (*N* = 43) | CHR (*N* = 40) | FEP (*N* = 70) |
|---|---|---|---|---|
| **Antipsychotics** | | | 6 (15%) | 63 (90%) |
| **Antidepressants** | | 2 (4.7%) | 8 (20%) | 15 (21.4%) |
| **Benzodiazepines** | | | 1 (2.5%) | 7 (10%) |
| **Anticholinergics** | | | | 4 (5.7%) |
| **Mood stabilizers** | | | 1 (2.5%) | 8 (11.4%) |
| **Other** | 7 (17.1%) | 2 (4.7%) | 2 (5%) | 9 (12.9%) |
| **Unmedicated** | 29 (70.7%) | 21 (48.8%) | 22 (55%) | 4 (5.7%) |
| **No information** | 5 (12.2%) | 18 (41.9%) | 2 (5%) | |

***Supplementary Table 3:*** self-reported medication usage by group. Note that antipsychotic use in the CHR group could largely be explained by low-dose seroquel (quetiapine) usage.

| | HC (*N* = 41) | FHR (*N* = 43) | CHR (*N* = 40) | FEP (*N* = 70) |
|---|---|---|---|---|
| **Alcohol** | 31 (75.6%) | 32 (74.4%) | 20 (50%) | 35 (50%) |
| **Amphetamines** | | 1 (2.3%) | | 1 (1.4%) |
| **Cannabis** | 9 (22%) | 13 (30.2%) | 12 (30%) | 19 (27.1%) |
| **Cocaine** | 1 (2.4%) | 3 (7%) | 2 (5%) | 3 (4.3%) |
| **Hallucinogens** | | 2 (4.7%) | 4 (10%) | 1 (1.4%) |
| **Sedatives/Hypnotics** | | 1 (2.3%) | 2 (5%) | |
| **Cigarettes** | 4 (9.8%) | 10 (23.3%) | 7 (17.5%) | 16 (22.9%) |
| **None** | 1 (2.4%) | 2 (4.7%) | 6 (15%) | 1 (1.4%) |
| **No information** | 2 (4.9%) | 6 (14%) | 11 (27.5%) | 14 (20%) |

***Supplementary Table 4:*** self-reported drug/substance use by group. No participants reported using inhalants, opioids, or PCP.

### 2. Symptoms

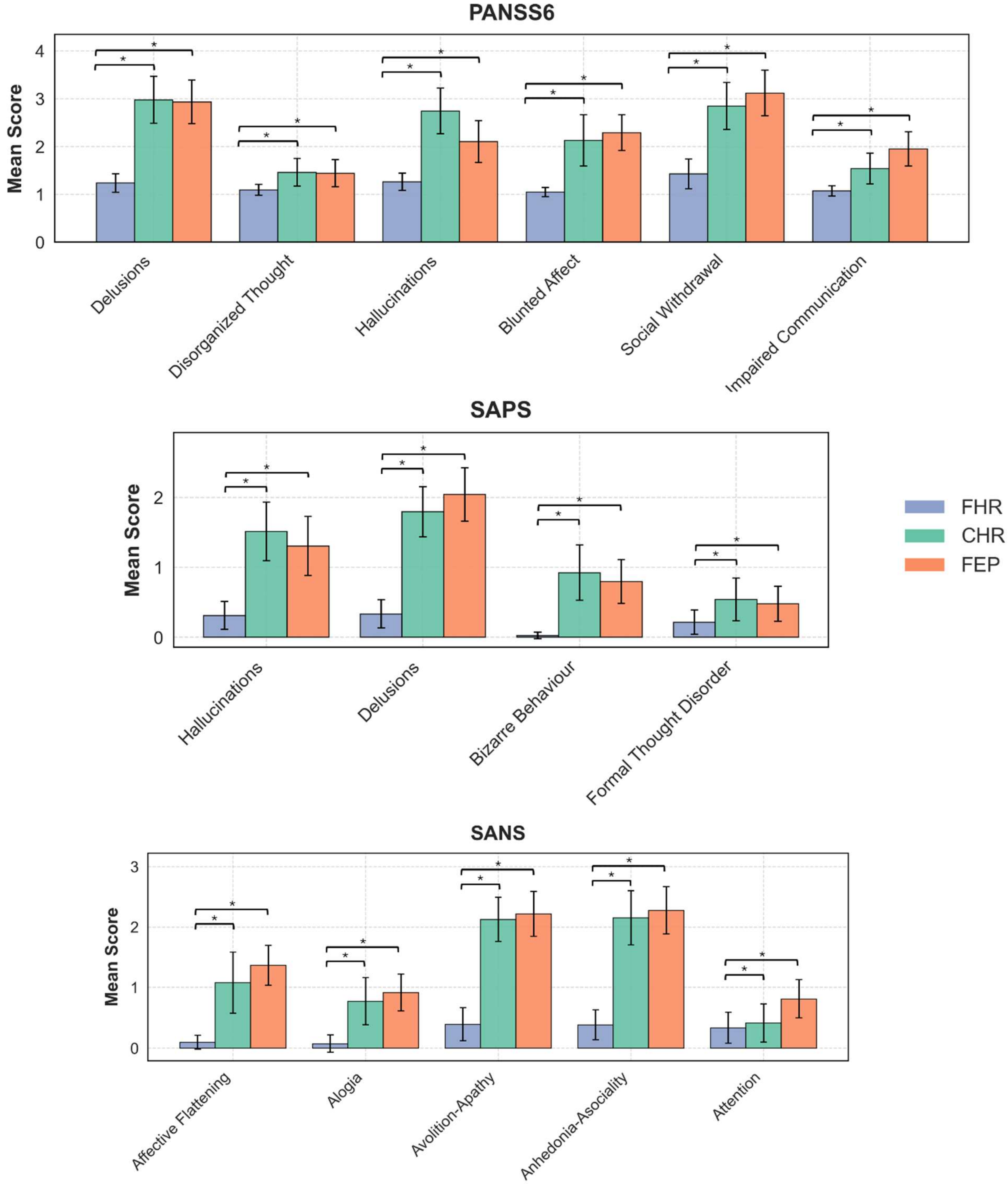


***Supplementary Figure 3:*** item-level group means and 95% confidence intervals for psychosis assessments. *$p < 0.05$ (uncorrected).

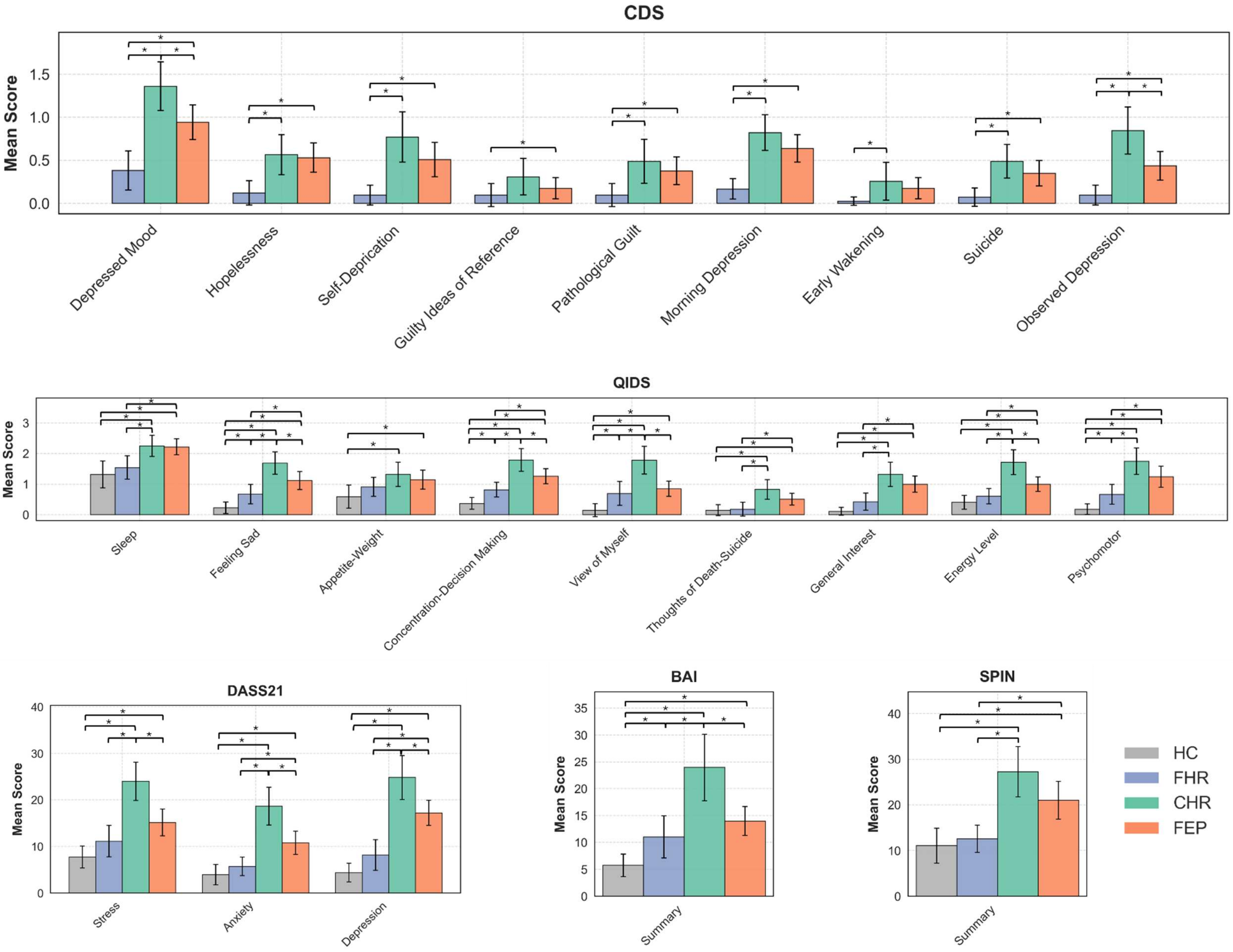


***Supplementary Figure 4:*** item-level group means and 95% confidence intervals for depression/anxiety assessments. *$p < 0.05$ (uncorrected).

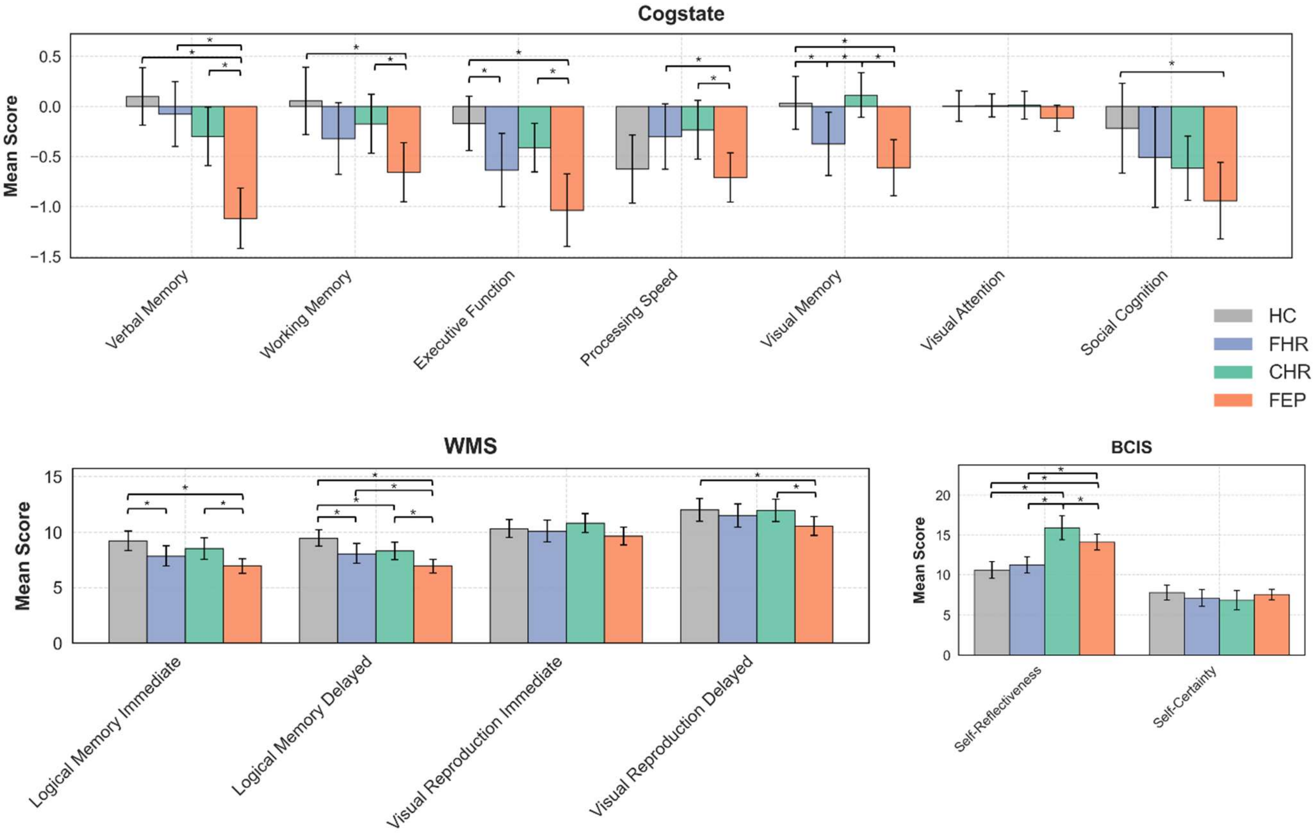


***Supplementary Figure 5**:* item-level group means and 95% confidence intervals for cognitive assessments. *$p < 0.05$ (uncorrected).

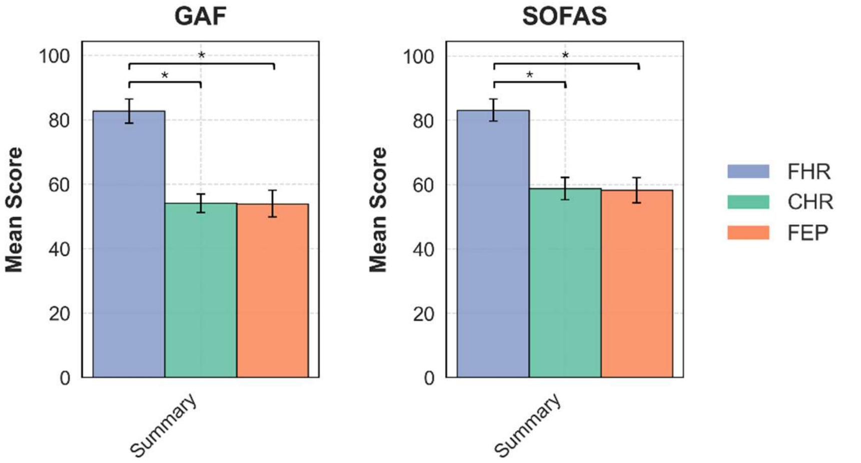


***Supplementary Figure 6**:* item-level group means and 95% confidence intervals for functioning assessments. *$p < 0.05$ (uncorrected).

### 3. Factor analysis

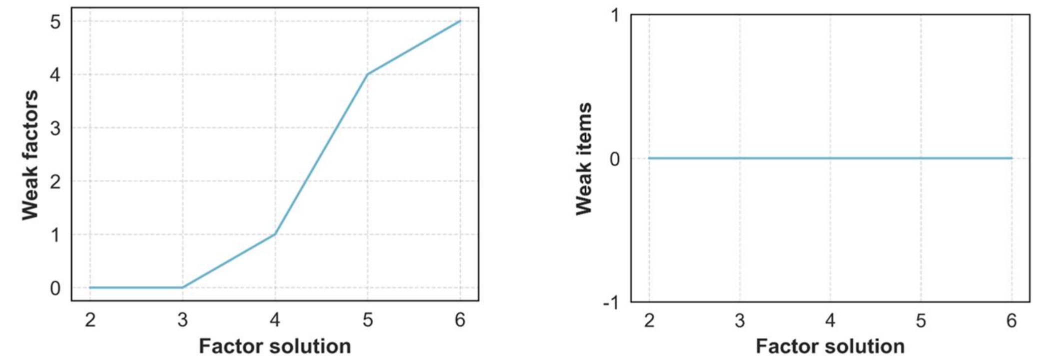


***Supplementary Figure 7:*** (*Left*) weak factors by solution, corresponding to the number of factors with less than 3 items with pattern loading > 0.5. (*Right*) weak items by solution, corresponding to the number of items with pattern loading < 0.3 on all factors. We chose 3 factors as the most granular solution which minimized both the number of weak items and factors.

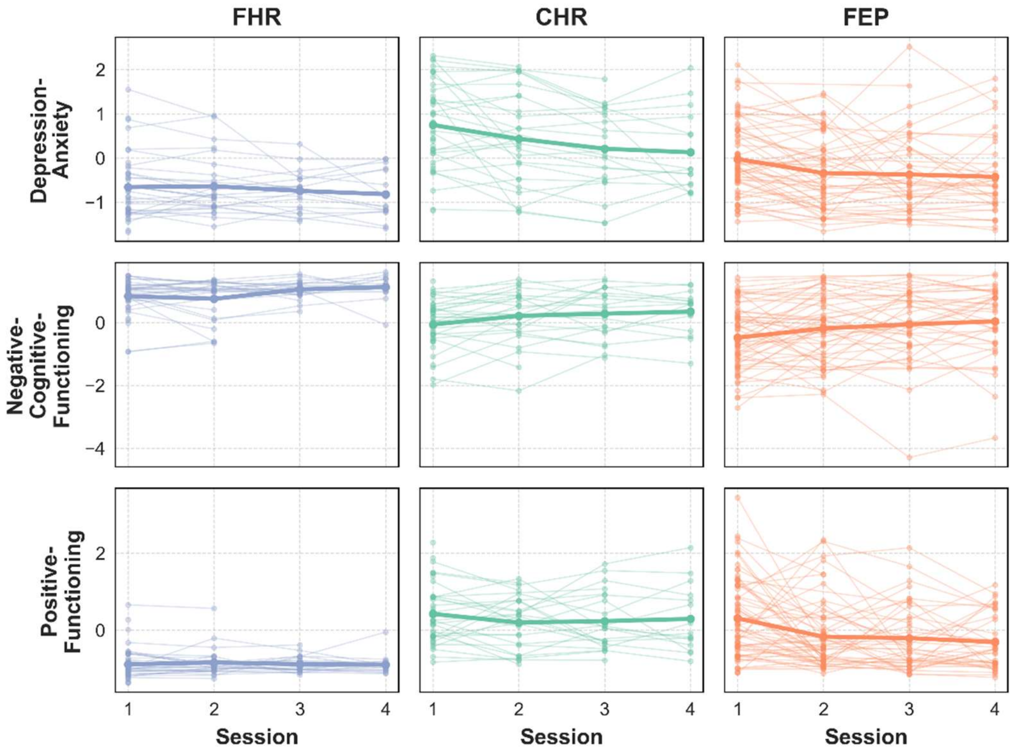


***Supplementary Figure 8:*** Longitudinal trajectories of factor scores for each group. Lighter lines connect observations from the same participants, while the thicker line indicates the timepoint-specific mean across participants. The interaction between the number of visits completed and time was not significant for any model (Depression-Anxiety: $p$ = 0.901; Negative-Cognitive-Functioning: $p$ = 0.689; Positive-Functioning: $p$ = 0.175), indicating that healthier participants at baseline did not necessarily return for more follow-up timepoints.

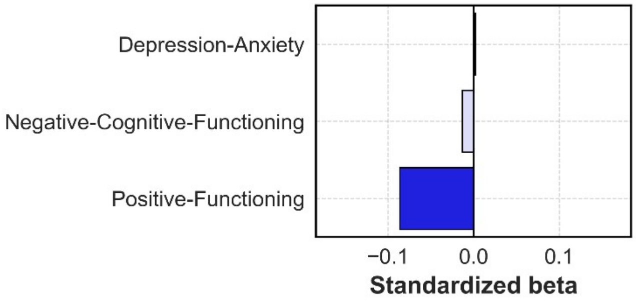


***Supplementary Figure 9:*** relationship between longitudinal factor scores and deviation from average medication dosage, covarying for average dosage, baseline age, sex, and time since baseline scan, allowing for random intercepts by participant. No effects were significant.

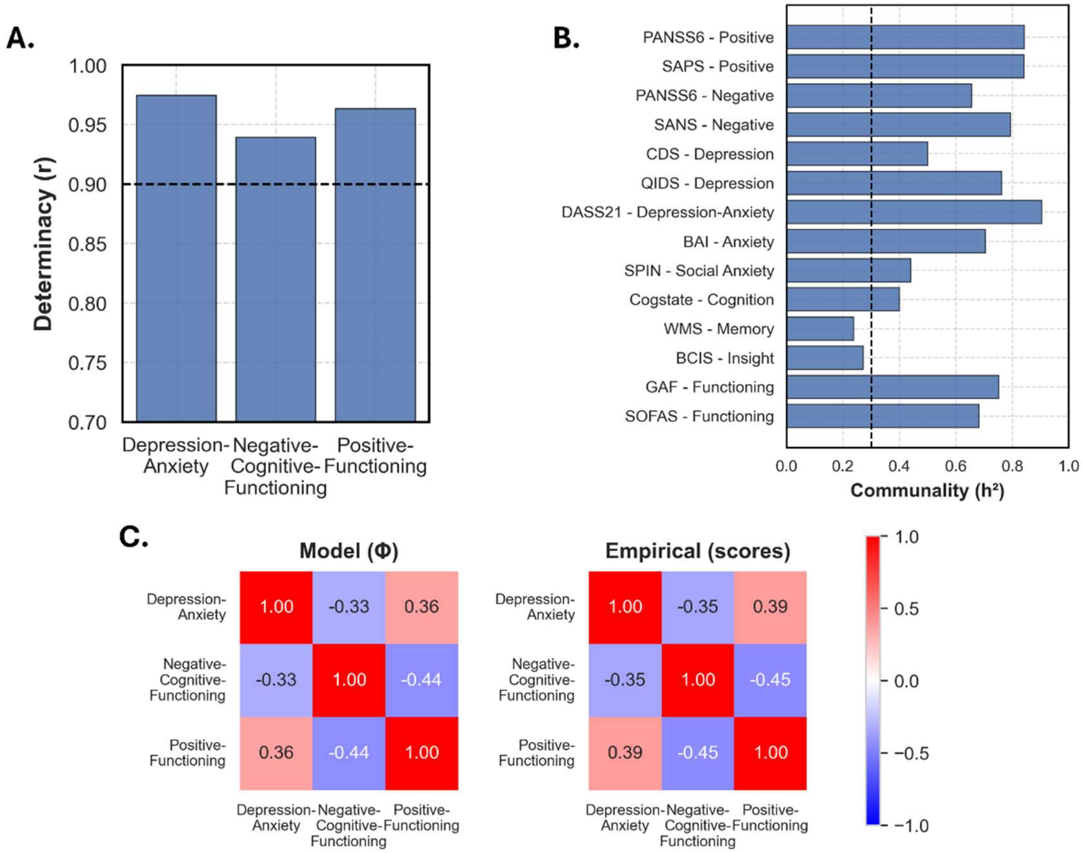


***Supplementary Figure 10:*** **(A)** Determinacy for each dimension. All factors were considered determinable, with values > 0.9. **(B)** Communalities for each variable, which were generally high, except for the WMS and BCIS. **(C)** Factor intercorrelations derived from the oblimin-rotated solution and correlations between ten Berge factor scores across participants. The factor scores successfully reproduced the correlation pattern.

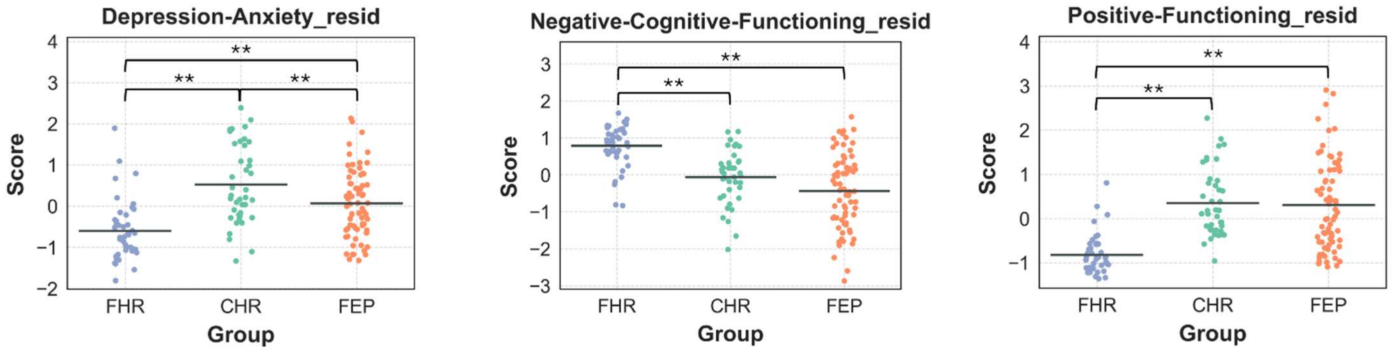


***Supplementary Figure 11**:* comparing factor scores between groups with additional covariates of age and sex. Results qualitatively replicated those shown in *Figure 1*, except for the trend towards significance between CHR and FEP on the negative-cognitive-functioning symptom cluster ($p$ = 0.063). ** $p_{Tukey} < 0.05$.

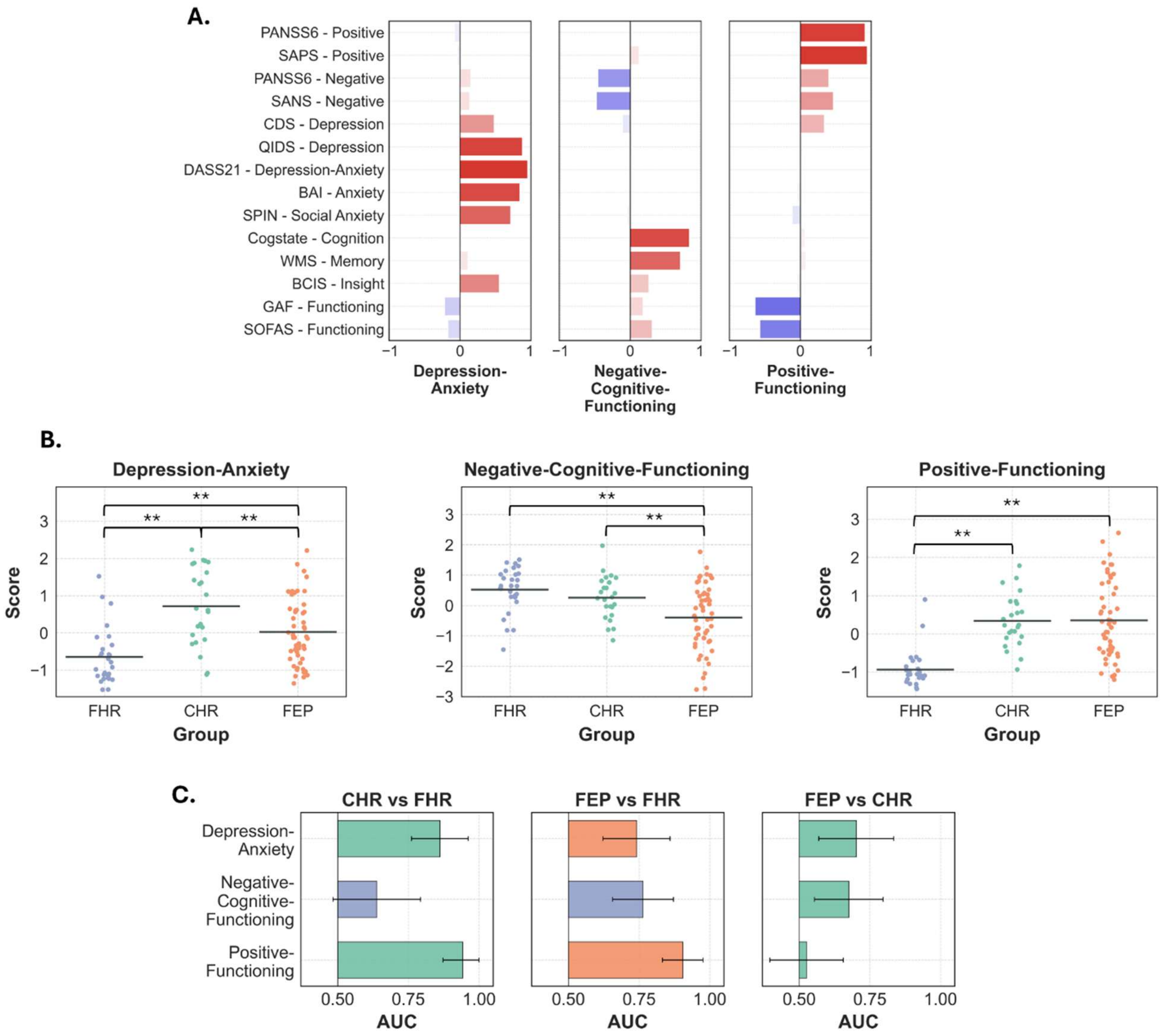


***Supplementary Figure 12**:* **(A)** Pattern loadings for the three factor solution in the subset of 104 participants who did not require multiple imputation at the summary scale level. Tucker congruence values indicated a highly consistent solution when compared to that shown in *Figure 1* (0.996, 0.933, and 0.943 for depression-anxiety, negative-cognitive-functioning, and positive-functioning, respectively). **(B)** Comparing factor scores between groups. The same pattern of results emerged as shown in *Figure 1*. $^{**}p_{Tukey} < 0.05$. **(C)** AUC values quantifying group overlap on each dimension, showing the same pattern of results as in *Figure 1*.

| | FHR (*N* = 43) | CHR (*N* = 40) | FEP (*N* = 70) |
|---|---|---|---|
| **PANSS-6** | | | |
| **SAPS** | | | |
| **SANS** | 1 (2.3%) | | 1 (1.4%) |
| **CDS** | | | 1 (1.4%) |
| **QIDS** | 2 (4.6%) | 2 (5%) | 16 (22.9%) |
| **DASS-21** | | | 1 (1.4%) |
| **BAI** | | | |
| **SPIN** | | | |
| **Cogstate** | 1 (2.3%) | | 1 (1.4%) |
| **WMS** | | | |
| **BCIS** | | | |
| **GAF** | | | |
| **SOFAS** | | | |

***Supplementary Table 5***: counts of participants missing individual observations for each scale. Missing values were replaced by their mean across the scale of interest.

| | FHR (*N* = 43) | CHR (*N* = 40) | FEP (*N* = 70) |
|---|---|---|---|
| **PANSS-6** | 1 (2.3%) | 1 (2.5%) | 11 (15.7%) |
| **SAPS** | 1 (2.3%) | 1 (2.5%) | 1 (1.4%) |
| **SANS** | 1 (2.3%) | 1 (2.5%) | 1 (1.4%) |
| **CDS** | 1 (2.3%) | 1 (2.5%) | 1 (1.4%) |
| **QIDS** | 8 (18.6%) | 11 (27.5%) | 14 (20%) |
| **DASS-21** | 9 (20.9%) | 11 (27.5%) | 15 (21.4%) |
| **BAI** | 10 (23.3%) | 13 (32.5%) | 15 (21.4%) |
| **SPIN** | 11 (25.6%) | 12 (30%) | 14 (20%) |
| **Cogstate** | 2 (4.6%) | | |
| **WMS** | 1 (2.3%) | 2 (2.5%) | 1 (1.4%) |
| **BCIS** | 11 (25.6%) | 11 (27.5%) | 4 (5.7%) |
| **GAF** | 1 (2.3%) | 1 (2.5%) | 1 (1.4%) |
| **SOFAS** | 1 (2.3%) | 1 (2.5%) | 5 (7.1%) |

***Supplementary Table 6:*** counts of participants missing all observations for a given scale. Missing values were replaced across 20 runs of multiple imputation.

### 4. Longitudinal analysis

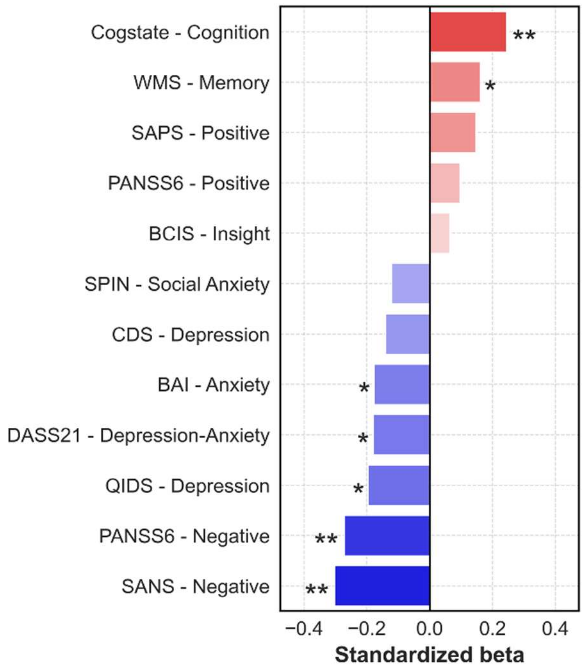


***Supplementary Figure 13:*** Standardized betas quantifying the effect of each assessment's scores on future global functioning. Each model was implemented separately and included covariates of baseline functioning, age, sex, group, and timepoint, along with a random effect of participant. Lower levels of negative symptoms, alongside greater levels of cognition, predicted better future functioning.

### 5. Neuroimaging analysis

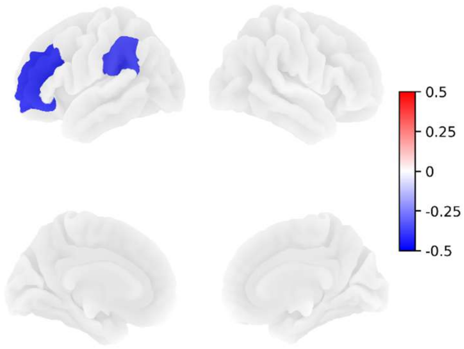


***Supplementary Figure 14:*** cortical thickness effect size map for FEP, thresholded at 10% false discovery rate. No regions passed at the same threshold for CHR or FHR.

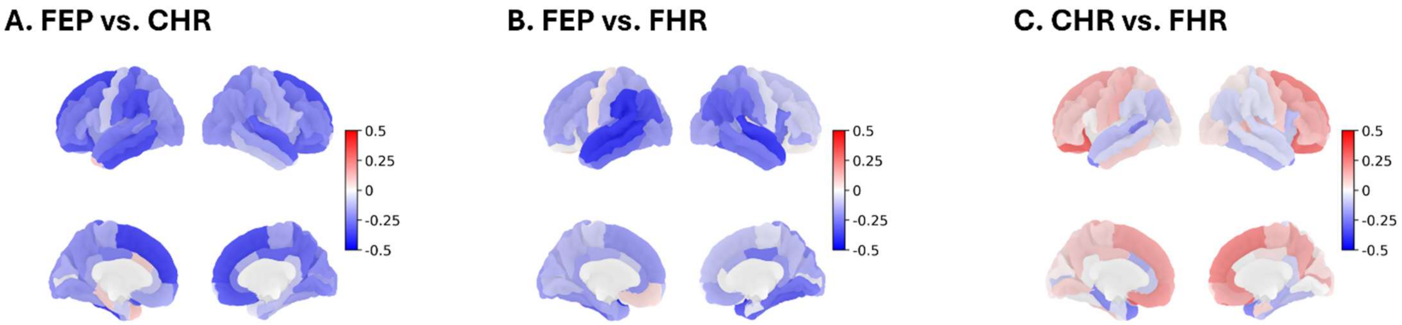


***Supplementary Figure 15:*** effect size maps showing regional effect of group on cortical thickness relative to non-control reference groups. The FEP vs. CHR ($r = 0.335$, $p_{spin} = 0.009$), FEP vs. FHR ($r = 0.472$, $p_{spin} < 0.001$), and CHR vs. FHR comparisons ($r = 0.643$, $p_{spin} < 0.001$) significantly correlated with the original comparisons between either group and controls, shown in *Figure 2C*.

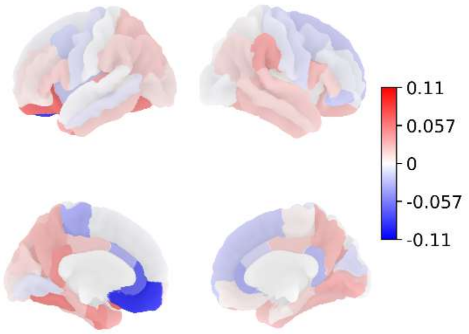


***Supplementary Figure 16:*** effect size map of deviation from average medication dosage on cortical thickness in the FEP group, covarying for average dosage, baseline age, sex, and time since baseline scan, allowing for random intercepts by participant. No regions survived at a 10% false discovery rate threshold. Qualitatively, medication was linked to a more spatially constrained pattern of cortical thickness reductions relative to the FEP map in *Figure 2*.

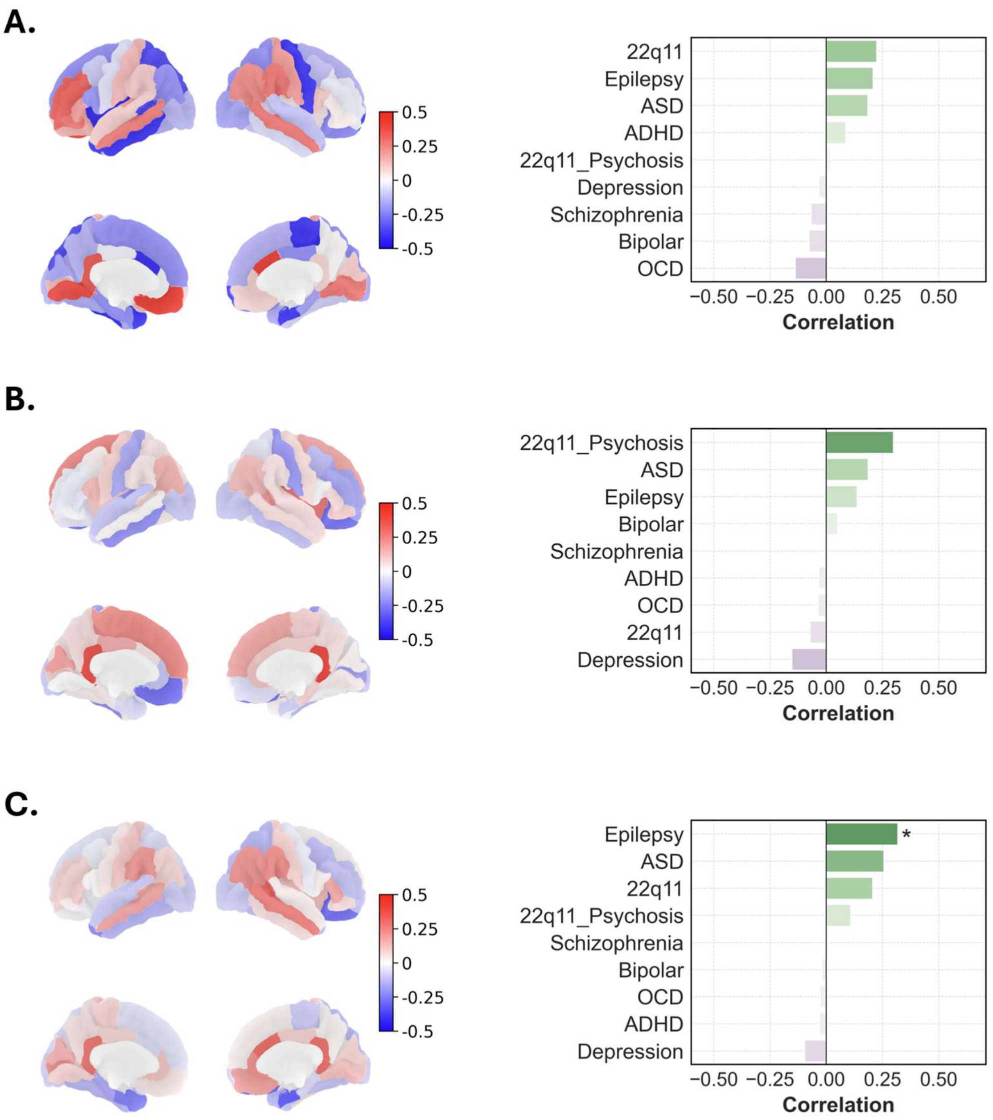


***Supplementary Figure 17:*** (*Left*) regional effect of group (Hedges' g) on cortical thickness relative to healthy controls while covarying for age, sex, and intracranial volume for CHR replication cohorts from London **(A)**, Mexico City **(B)**, and a combined map across both datasets **(C)**, additionally covarying for site. (*Right*) Spatial correlations between a given effect size map and comparable maps for 8 psychiatric, neurological, and developmental conditions provided by the ENIGMA toolbox. No consistent phenotype was identified across all three comparisons. **$p < 0.05$*

### 6. London CHR replication cohort

48 participants aged 18-40 were recruited through the South London and Maudsley NHS Trust, as well as public advertisement in the surrounding area, primarily to assess GABAergic receptor availability using positron emission tomography (PET). The sample included 24 antipsychotic-naive participants at clinical high-risk for psychosis with attenuated positive symptoms defined by the Comprehensive Assessment of At-Risk Mental States alongside 24 healthy controls. Exclusion criteria included any neurological condition, IQ < 70, PET or MRI contraindications, past or present substance abuse, pregnancy, and breastfeeding. Additionally, controls had no history of any mental health condition or family history of psychosis. All participants provided informed written consent for study participation. The study received ethics approval from the London/Surrey Research Ethics Committee. Sample demographics are summarized in *Table S3*.

T1-weighted images acquired using a 3T SignaTM PET-MR General Electric scanner. Images were processed and underwent quality control as described in the *Methods*, where 3 participants failed quality control. Effect size maps were generated as in the main text and compared with the ENIGMA maps accordingly.

| | HC | CHR | Statistic |
|---|---|---|---|
| **Age** (mean, std) | 25.2 (4.3) | 25.2 (4.8) | $t(46) = 0.019$,<br>$p = 0.985$ |
| **Male sex** (%) | 37.5 | 33.3 | $\chi^2(1,48) = 0$<br>$p = 1$ |
| **Neuroimaging quality control pass rate** (%) | 95.7 | 91.7 | $\chi^2(1,47) = 0$,<br>$p = 1$ |

***Supplementary Table 7:*** London CHR replication sample demographics. The first 4 columns show group-level summary statistics and the final column shows the results of an uncorrected t-test or chi-squared test assessing group differences in the statistic indicated. Comparisons were only performed on participants with the given measurement available.

### 7. Mexico City CHR replication cohort

151 participants aged 16-30 were recruited through the Instituto Nacional de Neurología y Neurocirugía in Mexico City. The sample included 84 participants at clinical high-risk for psychosis according to any SIPS criteria and 67 healthy controls (HC) with no history of psychiatric illness or family history of schizophrenia. Exclusion criteria included acute safety risk (e.g., suicidality), substance abuse within the previous month, substance dependence within the previous six months, substance use within one day of neuroimaging, any MRI contraindications (e.g., metallic implants), use of antipsychotic medication, a DSM-IV diagnosis of Bipolar I Disorder or Major Depression with Psychotic Features, severe claustrophobia, any significant organic brain disorder (e.g., epilepsy), pregnancy/lactation, and IQ < 70. All participants or their guardians provided informed written consent for study participation. The study was approved by a Research Ethics Board at the Instituto Nacional de Neurología y Neurocirugía. Sample demographics are summarized in *Table S4*

T1-weighted images were available for all but one participant. Participants were scanned on either a 3T Siemens Magnetom Skyra (MPRAGE) or a 3T General Electric Signa Excite (SPGR). Images were processed and underwent quality control as described in the *Methods*, where 2 participants failed quality control and another failed FreeSurfer processing. Effect size maps were generated as in the main text, with the scanner additionally included as a covariate, then compared with the ENIGMA maps accordingly.

| | HC | CHR | Statistic |
|---|---|---|---|
| **Age** (mean, std) | 22.2 (3.7) | 20.1 (4.1) | t(146) = 2.49, **p = 0.014** |
| **Male sex** (%) | 63.6 | 73.2 | $\chi^2$(1,148) = 1.14, p = 0.286 |
| **Years of education** (mean, std) | 14.8 (2.7) | 11.4 (3.3) | t(143) = 6.64, **p < 0.001** |
| **Neuroimaging quality control pass rate** (%) | 100 | 97.6 | $\chi^2$(1,150) = 0.32, p = 0.573 |
| **Siemens scanner** (%) | 41.8 | 26.2 | $\chi^2$(1,151) = 3.42, p = 0.064 |

***Supplementary Table 8:*** Mexico City CHR replication sample demographics. The first 4 columns show group-level summary statistics and the final column shows the results of an uncorrected t-test or chi-squared test assessing group differences in the statistic indicated. Comparisons were only performed on participants with the given measurement available.

### 8. Partial least squares analysis

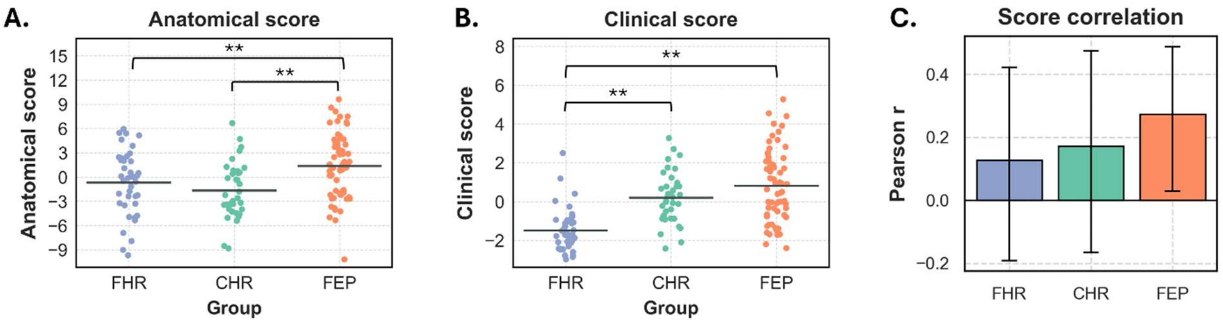


***Supplementary Figure 18:*** PLS scores for anatomical **(A)** and clinical **(B)** components of the pattern shown in *Figure 3.* FEP only expressed the anatomical, and not the clinical, pattern more than the other groups. **(C)** Correlations between scores for each group. The correlation was only significant for FEP, though the correlations did not significantly differ from each other.

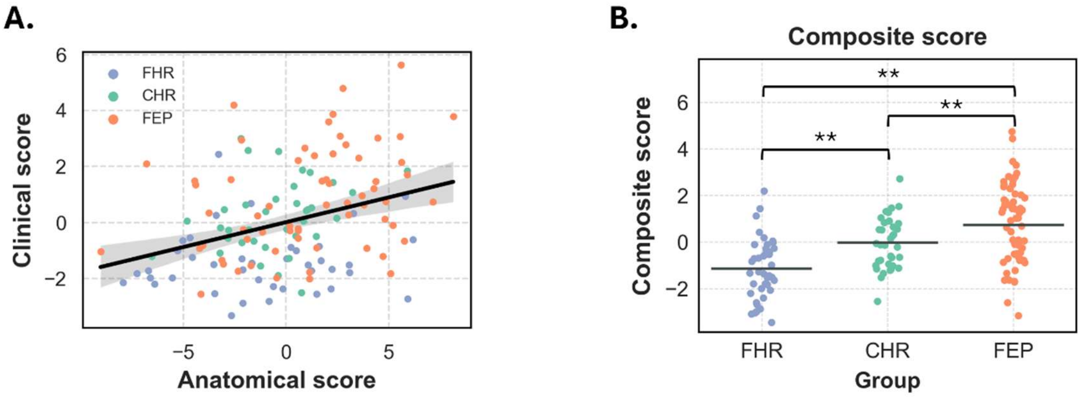


***Supplementary Figure 19:*** Correlations between PLS scores ($r$ = 0.344, $p$ < 0.001) **(A)**, alongside comparisons of composite scores by group ($F$(2,137) = 20.63, $p$ < 0.001) **(B)**, after residualizing for age and sex. Results replicate those shown in *Figure 3C-D*.

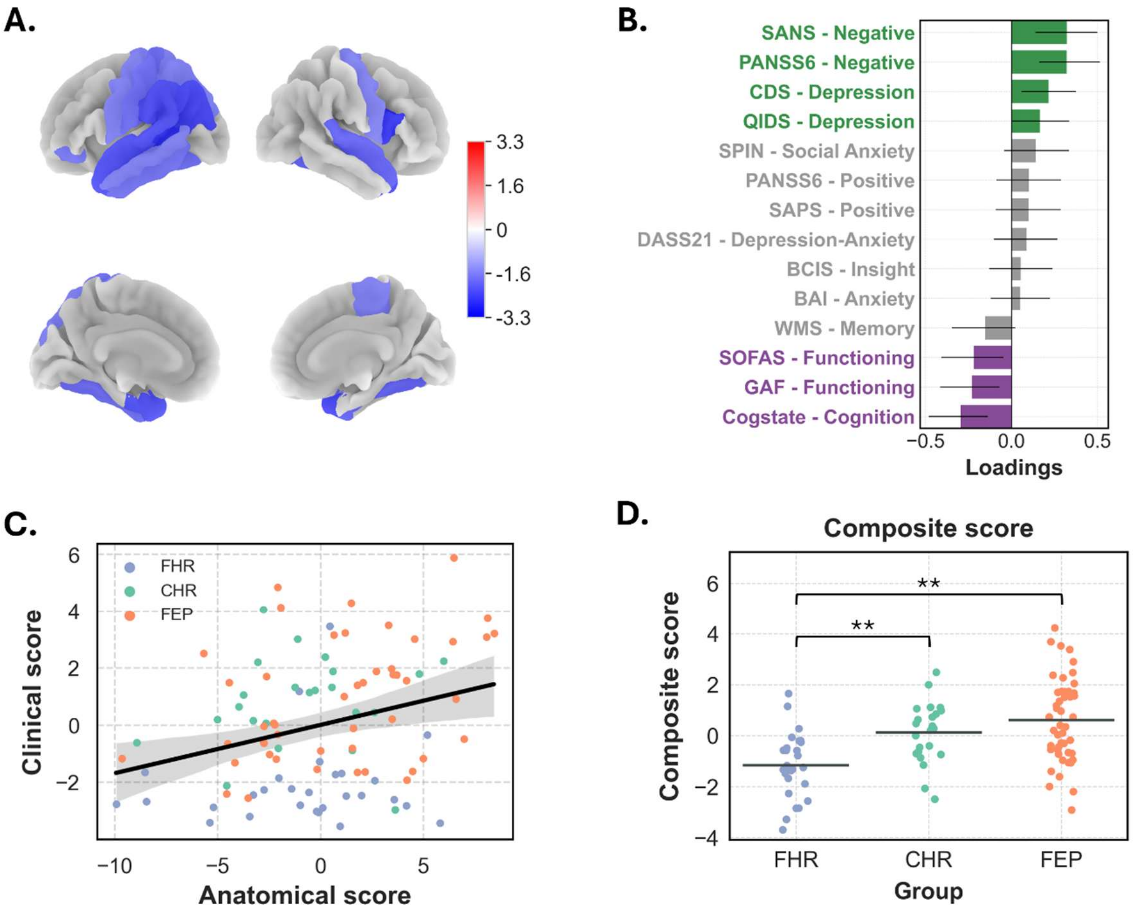


***Supplementary Figure 20:*** The first latent variable from our partial least squares analysis performed on a smaller subsample (*N* = 99) who did not require multiple imputation at the summary scale level. Tucker congruence values indicated that the anatomical (0.946) and clinical (0.921) loadings were highly consistent with those shown in *Figure 3A-B*. Note that FEP no longer expressed the composite score more than the other groups in this smaller subsample, though this analysis was only intended to assess the replicability of the dominant brain-behaviour pattern without data imputation.
